# Integrating NMR Metabolomics and Glycomics for Early Cancer Detection in Patients with Non-Specific Symptoms

**DOI:** 10.64898/2026.01.20.26344357

**Authors:** Tereza Kacerova, Abi G. Yates, James R. Larkin, Boris Shulgin, Jack J. J. J. Miller, Philippa L. Harris Gleave, Sebastian de Jel, Jack Cheeseman, Georgia Elgood-Hunt, the SCAN Consortium, Eric Schiffer, Daniel I. R. Spencer, Suzie Anthony, Daniel C. Anthony

## Abstract

**Background:** Early cancer diagnosis in patients with non-specific symptoms is limited by the lack of discriminatory tests. Within the Oxfordshire Suspected CANcer (SCAN) pathway, exploratory biomarker work showed that serum ^1^H-NMR-based metabolomics can identify cancer with high accuracy. SCAN2 tested whether integrating metabolomics with glycomics improves discrimination in a clinically complex, real-world population.

**Methods:** Serum from 369 SCAN patients (59 cancers) was analysed using *AXINON®lipoFIT®*-derived NMR metabolomics and HPLC-MS glycomics. Machine-learning models were trained to predict cancer status, with performance assessed by receiver operating characteristic (ROC) analysis of pooled cross-validated predictions. To place cancer risk in a broader clinical context, a second classifier modelling alternative non-cancer diagnosis was incorporated, and mean predicted probabilities from both models were jointly projected into a two-dimensional space, maintaining strict separation of training and test data.

**Findings:** Integration of glycomics with metabolomics improved discrimination, achieving an AUC of 0.88 in a refined cohort excluding dominant comorbidities. Cancer-associated bi- and tri-antennary glycans, including FA2G2S1, FA2BG1, and M5A1G1S1, differentiated cancer cases. A classifier targeting metastatic disease achieved an AUC of 0.80. Joint probability analysis preserved cancer-associated metabolic signatures across comorbidity burden, with projection-based classification achieving an accuracy of 89.8%.

**Interpretation:** These findings validate the SCAN1 metabolomic signature in a more clinically complex cohort and demonstrate that integrating metabolomics with glycomics enhances cancer detection in patients with non-specific symptoms. Joint probability analysis provides an interpretable framework for cancer risk stratification within multimorbid diagnostic pathways, supporting the clinical potential of scalable multi-omics blood testing.

## INTRODUCTION

Stage at diagnosis remains a key determinant of cancer outcomes. While current referral pathways from primary care to site-specific referral pathways perform well for patients with localising symptoms, they are less effective for individuals presenting with non-specific symptoms such as fatigue, weight loss, or persistent pain.^1–3^ For these patients, lack of a dedicated referral pathway frequently results in delayed clinical assessment and a higher likelihood of advanced disease at presentation.^3,4^

Multidisciplinary diagnostic centre (MDC) models were developed to address diagnostic gaps in patients with non-specific symptoms and have been associated with improved early cancer detection^5^ and identification of less common malignancies.^6^ In England, this approach was formalised through the Accelerate, Coordinate, Evaluate (ACE) programme and the subsequent implementation of Rapid Diagnostic Centres (RDCs).^7^ The Oxford Suspected CANcer (SCAN) pathway, initially established as an ACE Wave 2 pilot and now part of standard care, provides a real-world framework for evaluating diagnostic strategies in primary care referrals with non-specific symptoms.^8^ However, despite improved access to investigation, referral to non-site-specific pathways remains largely reliant on primary care clinical judgement, resulting in heterogeneity in referral thresholds and timing.^9,10^

Beyond imaging and routine blood tests, blood-based molecular profiling has emerged as a potential tool for earlier risk stratification in patients presenting with non-specific symptoms, reflecting the systemic metabolic impact of cancer.^11^ Metabolomic approaches have been widely applied in preclinical models and in clinically defined, site-specific malignancies, but remain underexplored in non-site-specific referral populations.^12^

In the initial SCAN study (SCAN1), blood metabolomic profiling using nuclear magnetic resonance (NMR) spectroscopy enabled discrimination between patients with non-specific symptoms with and without malignancy, and further stratified malignant cases according to metastatic status, despite substantial underlying clinical heterogeneity.^13^ Diagnostic performance was high, with sensitivities and specificities exceeding 80% for solid tumours and comparable performance for metastatic disease.^14^ A shared metabolic signature across tumour types, which was dominated by lipid-associated resonances and inflammatory glycoprotein signals, suggested systemic metabolic reprogramming rather than site-specific effects. The findings supported metabolomics as a biologically grounded triage tool within non-site-specific diagnostic pathways, complementing clinical judgement by prioritising investigation in higher-risk patients while safely excluding malignancy in others. However, limitations remained: SCAN1 was modest in size, pre-analytical handling reflected routine practice, and metabolomics alone captures only a partial host response to cancer.

Tumour-associated inflammation and altered protein glycosylation are established features of malignancy but are only partially captured by standard NMR metabolomic profiles.^15,16^ In SCAN1, the prominence of *N*-acetylated glycoprotein and sialic acid signals (GlycA/GlycB) within the cancer-associated metabolic signature indicated a contribution from inflammatory glycoprotein remodelling that could not be fully resolved by NMR alone, motivating the integration of targeted glycomic profiling to assess its complementary diagnostic value.^16,17^ The SCAN2 study was therefore designed to evaluate an expanded multi-omics framework in a larger, clinically more complex population referred through the routine SCAN pathway. Stricter pre-analytical standardisation of samples was implemented, and untargeted and targeted NMR metabolomics were combined with liquid chromatography-mass spectrometry (LC-MS)-based glycomic profiling to enable concurrent assessment of small-molecule metabolites, lipoprotein subfractions, and *N*-acetylated glycoproteins.

Here, we show that the core findings of SCAN1 are validated by SCAN2 despite greater clinical complexity, and that inclusion of glycomic features improves diagnostic performance. These results support integrated blood-based multi-omics profiling as a complementary approach within MDC and RDC pathways for cancer detection and metastatic risk stratification in patients with non-specific symptoms.

## MATERIALS AND METHODS

### Patient recruitment and eligibility criteria

Patients were recruited from the Oxfordshire Suspected CANcer (SCAN) pathway between November 2020 and July 2021, using the same non-site-specific referral framework described in the original SCAN study.^14,18^ Adults aged over forty years were eligible if referred from primary care with one or more non-specific but concerning features suggestive of cancer or other serious disease. These included unexplained weight loss, severe fatigue, persistent nausea or anorexia, new or atypical pain without a clear site-specific cause, unexplained laboratory abnormalities (e.g. anaemia, thrombocytopenia, hypercalcaemia), or general practitioner (GP) clinical suspicion of serious disease. The latter captured cases in which the overall symptom constellation, despite being non-specific, was judged sufficiently concerning to warrant urgent investigation. Collectively, these criteria reflect the role of the SCAN pathway as a diagnostic route for patients without organ-specific symptoms but at appreciable risk of malignancy. The primary analytical model included all recruited participants. As SCAN1 demonstrated superior metabolomic discrimination for solid-organ compared with haematological malignancies, a predefined subgroup analysis in SCAN2 excluded patients with haematological cancers. To further optimise discriminatory performance, individuals with uncertain or non-biopsy-confirmed cancer diagnoses were excluded, including those with pre-malignant conditions such as Barrett’s oesophagitis and monoclonal gammopathy of undetermined significance (MGUS). To minimise metabolic confounding, patients with significant renal impairment were excluded given the known effects of renal dysfunction on circulating lipid and glycoprotein profiles. Participants with prostate cancer were also excluded, as androgen-deprivation therapy substantially alters systemic metabolism and information on active or recent treatment was unavailable.

Written informed consent was obtained from all participants. The study was conducted in accordance with the UK Policy Framework for Health and Social Care Research and received ethical approval from the Oxford Radcliffe Biobank research tissue bank (20/A120).

### Pre analytical sample handling and blood collection

Blood samples for metabolomic analysis were collected immediately prior to CT imaging using standardised pre-analytical procedures.^19^ Venous blood was drawn into serum tubes without additives, following a mandated minimum fasting period of two hours. Tubes were gently inverted and allowed to clot at ambient temperature for a standardised interval before centrifugation at 2,200 g for 10 minutes within 30-60 minutes of collection. Serum was inspected for haemolysis, aliquoted into low-binding polypropylene vials, and transferred to dry ice within minutes of centrifugation before storage at -80 °C. All collection times, processing intervals, and protocol deviations were recorded prospectively.

### NMR metabolomics acquisition

Serum aliquots were thawed at 4 °C and inspected visually for evidence of protein precipitation prior to analysis. Samples were prepared using a standardised protocol implemented by Numares AG (Regensburg, Germany), involving dilution with a deuterated phosphate buffer to ensure chemical shift stability and field locking.^20^ Serum was transferred to 5 mm NMR tubes for spectral acquisition. All ^1^H NMR experiments were performed at Numares AG using a 600 MHz Bruker AVIII spectrometer. One-dimensional spectra were acquired using a presaturation-based pulse sequence to attenuate the residual water signal and were processed using proprietary software developed by Numares AG.^20,21^ In addition to targeted, quantitatively calibrated metabolite and lipoprotein parameters derived using the *AXINON®lipoFIT®* NMR platform through spectral deconvolution, untargeted metabolomic features were generated by segmenting spectral regions between 0.55-4.25 ppm and 5.20-8.50 ppm into fixed-width 0.01-ppm bins.^22^ Spectra were referenced to the lactate methyl doublet (δ = 1.33 ppm). Untargeted metabolite assignment was supported by reference to previous experiments, literature,^19,23^ and the Human Metabolome Database.^24^

### HPLC-HRMS *N*-Glycomic profiling

Serum *N*-glycan profiling was performed by Ludger Ltd. (Oxford, UK) using established and previously published workflows.^17^ *N*-glycans were enzymatically released from glycoproteins using peptide-*N*-glycosidase F (PNGase F), fluorescently labelled, and analysed by hydrophilic interaction liquid chromatography coupled to high-resolution mass spectrometry with fluorescence detection (HILIC-HRMS). Chromatographic separation was performed on a BEH-Glycan column using a Vanquish UHPLC system (Thermo Fisher Scientific), coupled to an Orbitrap Exploris 120 mass spectrometer operated in positive ion mode. System suitability standards and blanks were included throughout the analytical run to monitor performance and stability. Glycomics data were processed in Skyline software. Both fluorescence-derived chromatographic peaks, which typically represent composite glycan groups, and mass spectrometry-derived peaks corresponding to individual assigned glycans were quantified and analysed, alongside models combining both feature types. Full details of sample preparation, chromatographic gradients, and mass spectrometric parameters have been described previously.^17^

### *N*-Glycan Nomenclature

Glycan structures were described using the standard Oxford Glycan Notation.^25^ *N*-glycans are covalently attached oligosaccharides characterised by a conserved pentasaccharide core composed of two *N*-acetylglucosamine (GlcNAc) and three mannose residues, which is subsequently elaborated through the enzymatic addition of specific monosaccharides, including fucose (**F**), additional mannose (**M**), galactose (**G**), and sialic acid (**S**), as well as by bisecting *N*-acetylglucosamine (**B**) and variations in branching patterns (antennae; **A**).^25^ Numeric descriptors indicate the number of each residue present, allowing concise representation of glycan composition.

### Multivariate statistical analysis

To evaluate differences between groups, we developed an *in-house* analysis framework implemented in R (v4.5.2), referred to as the BTK model. Missing features were addressed using sample-wise *k*-nearest neighbours (kNN) imputation. NMR data were sum normalised and the glycomics data were median normalised and log transformed prior to statistical analysis. All input features were subsequently *z*-scored to harmonise scales across measurements. Group discrimination was assessed using random forest classifiers with embedded feature selection and repeated stratified *k*-fold cross-validation. In each iteration, folds were constructed by assigning four cancer cases per fold, resulting in *k* = 14 folds (⌊59/4⌋), with class stratification preserved across splits. Model performance was evaluated using receiver operating characteristic (ROC) analysis on pooled out-of-fold predictions.^26,27^ An initial random forest model was used to rank variables by mean decrease in Gini index, with the highest-ranking features retained for downstream modelling. The cross-validation procedure was repeated across 100 independent random permutations to assess model stability and reduce sensitivity to data partitioning. Within each cross-validation iteration, feature importance was recalculated using training data only, and class membership probabilities were generated for held-out test samples.^26,28^ ROC curves were constructed by varying the classification threshold across the full range of predicted probabilities. Sensitivity (true positive rate), specificity (true negative rate), and overall accuracy were computed at each threshold by comparing predicted class labels with observed outcomes, yielding corresponding counts of true positives (TP), true negatives (TN), false positives (FP), and false negatives (FN). Threshold-dependent performance metrics were averaged across resampling iterations. Feature rankings were obtained by averaging variable importance scores across cross-validation repeats, and a reduced feature set was selected to minimise overfitting. Training and test data were separated within each iteration. Model robustness was assessed using standard label-randomisation procedures.

### BTK dual-outcome prediction framework

To jointly characterise cancer status and comorbidity burden, the BTK framework was extended to a dual-outcome modelling strategy. Molecular feature selection was performed exclusively within the cancer-focused subgroup defined above, and the resulting reduced feature set was fixed for all subsequent analyses. Using this shared feature space, an independent random forest model was then trained to distinguish alternative non-cancer diagnoses from healthy controls. Individuals not meeting the training definition for the alternative non-cancer diagnosis model were not used for model fitting but were instead projected into the trained model to obtain probabilistic estimates. This design ensured that both outcomes were interrogated within a common molecular feature space while preserving statistical independence between models. Cancer and alternative non-cancer diagnosis models were trained separately within each cross-validation fold, with class probabilities generated exclusively for held-out samples. For each individual, predicted probabilities were averaged across all test-fold occurrences to obtain stable estimates of cancer and alternative non-cancer diagnosis risk. Model performance was assessed using ROC analysis based on pooled cross-validated predictions.

To examine the joint structure of these predictions, individuals were projected into a two-dimensional space defined by the mean predicted probability of cancer and the mean predicted probability of an alternative non-cancer diagnosis, represented on the logit scale.^29,30^ The distribution of samples within this joint probability space was first summarised using two-dimensional Gaussian kernel density estimation (KDE), with iso-density contours enclosing a fixed proportion of the probability mass.^29^ This approach provides a threshold-free, data-driven description of how individuals populate the joint cancer-alternative non-cancer diagnosis risk landscape, without imposing predefined decision boundaries. For downstream classification and performance evaluation, explicit geometric decision regions were defined within the joint cancer-alternative non-cancer diagnosis probability space. The distribution of non-cancer individuals was characterised by an elliptical envelope in the model-derived probability space, defined by the empirical mean vector and covariance matrix of the non-cancer class. This envelope captured the dominant region of multivariate probability mass associated with non-cancer profiles. In parallel, a directional wedge-shaped region was defined to capture the cancer-associated probability signal, with fixed orientation and data-driven scaling anchored to the region of highest cancer-positive prediction density. Individuals falling within the wedge were classified as predicted cancer-positive, enabling construction of confusion matrices and calculation of classification metrics. Importantly, KDE was used exclusively for descriptive visualisation of the joint probability structure, whereas the wedge-based decision region was introduced solely to convert the continuous risk space into a discrete classification framework. All analyses were conducted with strict separation of training and test data.

### Univariate statistical analysis

Univariate associations between molecular traits and diagnostic group were assessed using odds ratios (ORs estimated from binary classification models implemented in R (v4.5.2)), quantifying the change in diagnosis probability per unit increase in each feature. For each multivariable model, univariate analyses were restricted to the top 30 features identified by mean decrease in Gini index from the BTK random forest feature-ranking step. Results were visualised as forest plots with 95% confidence intervals. For continuous variables, group differences were evaluated using unpaired two-sample *t* tests with false discovery rate correction (Benjamini-Hochberg, FDR < 0.05), while categorical variables were compared using the chi-square test. Correlations among significantly altered molecular features were assessed using Pearson correlation analysis. Data are presented as Tukey boxplots with whiskers extending to 1.5× the interquartile range (GraphPad Prism 10).

## RESULTS

### Cohort characteristics and application of eligibility criteria

The SCAN2 cohort comprised 369 primary care referrals with non-specific symptoms, of whom 59 were diagnosed with cancer and 310 had malignancy excluded. Recruitment was conducted under stricter pre-analytical conditions than SCAN1, including standardised fasting and immediate serum separation, to reduce technical variability. Despite these refinements, the cohort remained clinically heterogeneous, with diverse tumour sites and multiple comorbidities across diagnostic group.

Three complementary analytical models were constructed (Fig. 1). The primary model included all participants (n = 369) and assessed overall discrimination between cancer (n = 59) and non-cancer cases (n = 310). The second model specifically targeted solid-organ cancers, excluding patients with haematological malignancies. In addition, in this cohort we excluded patients with a diagnosis of a pre-malignant condition or where the diagnosis of cancer was non-biopsy proven, patients with prostate cancer, and patient with renal impairment (see methodology). This group comprised 32 cancer cases and 277 non-cancer participants (Fig. 1). Finally, to examine whether metabolic and glycomic signatures varied with disease burden, a third model stratified all cancer cases in the cohort into metastatic (n = 29) and non-metastatic (n = 30) groups.

**Figure 1.**
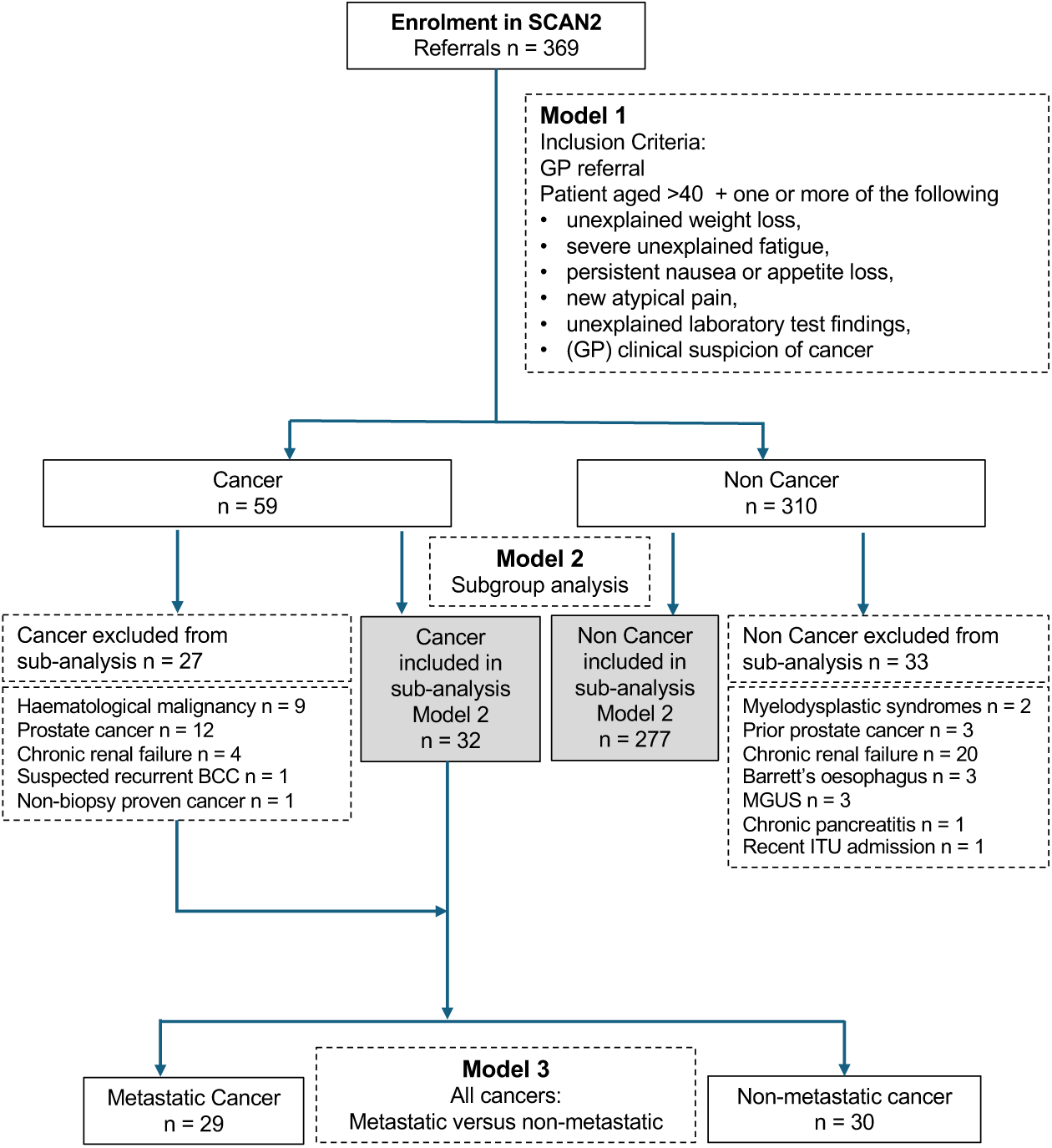
Overview of the SCAN2 cohort and study inclusion criteria. Flow diagram summarising participant selection from the SCAN pathway, key inclusion and exclusion criteria, and subgroup definitions used for cancer and non-cancer comparisons. *MGUS:* Monoclonal gammopathy of undetermined significance, *BCC:* basal cell carcinoma, *ITU:* intensive therapy unit.

Demographic and clinical characteristics of participants across the three analytical models are summarised in Table 1. The SCAN2 cohort comprised 369 individuals (59 cancer, 310 non-cancer), with a broad range of cancer types represented (SI Table 1). This heterogeneity reflects the non-site-specific diagnostic setting of the study. Across the entire cohort, participants with cancer were significantly older than non-cancer participants (74.7 ± 10.1 vs. 69.5 ± 13.0 years; p = 0.004), whereas sex distribution and body mass index (BMI) did not differ between groups. A similar age difference was observed in the rigorously stratified subgroup model (75.0 ± 10.8 vs. 68.3 ± 12.9 years; p = 0.005), again with no significant differences in sex or BMI. In contrast, no significant differences in age, sex, or BMI were observed between metastatic and non-metastatic cancer cases.

**Table 1.**
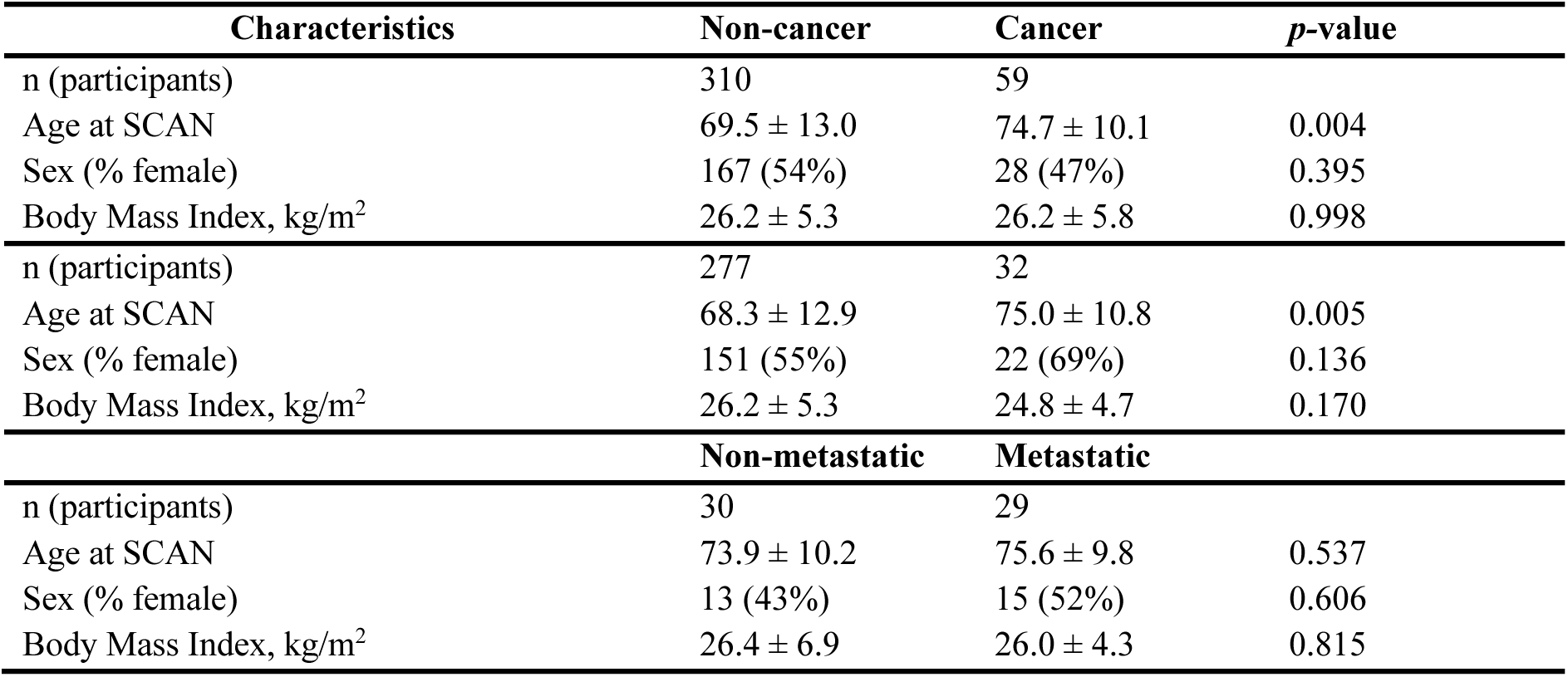
Demographic and clinical characteristics of patients across the three classification models. (all SCAN2 participants, rigorously stratified sub-cohort, and metastatic cancer model). Data are presented as mean ± standard deviation. Between-group comparisons were performed using unpaired two-tailed t-tests for continuous variables and chi-square tests for categorical variables. A p-value < 0.05 was considered statistically significant. BMI data were available for 340 of 369 patients in the full cohort, for 288 of 309 patients in the rigorously stratified model, and for 54 of 59 patients in the metastatic cancer model.

The observed age difference between cancer and non-cancer participants likely reflects the larger and more heterogeneous non-cancer cohort. To evaluate the impact of age on model performance, classification models were constructed with and without age as a covariate. Inclusion of age yielded an ROC AUC of 0.814 (95% CI 0.808-0.820), while exclusion resulted in a comparable AUC of 0.815 (95% CI 0.809-0.821), indicating minimal influence of age on discrimination. Consistent with this, age ranked 258th in variables by feature importance and was not included in the primary predictive signature. Correlation analyses between age and top-ranked features showed uniformly weak associations (|r| < 0.35), confirming that model performance was driven by metabolic and glycomic features rather than age-related effects (SI Figs. 1-4). Additionally, estimated glomerular filtration rate (eGFR), determined from serum creatinine levels, showed a modest univariable association with cancer status (p = 0.013, SI Fig. 5A), but it was not prioritised in multivariable modelling, ranking 106th in variables by feature importance, and its inclusion did not materially affect model performance (ROC AUC 0.809 (95% CI 0.803-0.815)), indicating that residual renal function differences did not drive discrimination (SI Fig. 5B).

### Replication of the SCAN1 metabolomic signature in a more heterogeneous cohort

We previously reported that serum metabolomic profiling robustly discriminated cancer from non-cancer participants, and metastatic from non-metastatic disease, in the SCAN1 cohort. To evaluate the generalisability of these findings under increased clinical heterogeneity, we performed independent metabolomic profiling and model development in the SCAN2 cohort using a different analytical NMR FDA-cleared platform and modelling strategy.

In SCAN2, multivariable modelling identified nine NMR-derived metabolomic features (glutamate, glutamine, histidine, myo-inositol, an overlapping valine/proline signal, alanine, lactate, sialic acid signal (GlycB), and high-density lipoproteins (HDLs)) that were able to discriminate cancer from non-cancer participants with a ROC AUC of 0.78 (SI Fig. 6A). Predicted probability distributions showed clear separation between diagnostic groups, indicating preservation of classification structure despite substantial cohort heterogeneity (SI Fig. 6B).

To assess external validity and exclude cohort- or platform-specific effects, the SCAN2-derived metabolite panel was then applied to the previously published SCAN1 cohort. This reciprocal validation yielded stronger discrimination (AUC 0.88; SI Fig. 6C), with predicted versus observed probabilities confirming robust separation between cancer and non-cancer participants (SI Fig. 6D). Univariate odds ratios were directionally concordant (SI Fig. 6E), demonstrating consistency of individual metabolite-disease associations.

Together, these findings demonstrate bidirectional replication across temporally independent cohorts, analytical platforms, and modelling strategies. Discriminatory metabolic structure originally reported in SCAN1 was reproduced in the more heterogeneous SCAN2 cohort, while metabolites independently prioritised in SCAN2 retained both discriminatory performance and effect direction when re-evaluated in the previously published SCAN1 dataset. This convergence provides strong evidence for the robustness and generalisability of the NMR-derived metabolic signature in non-specific symptom populations.

### NMR metabolomics and LC-MS glycomics detect cancer in a cohort with non-specific symptoms

To enhance performance in the SCAN2 cohort and better capture lipid-related metabolic perturbations, the NMR-only framework was extended to incorporate deconvoluted, lipoprotein-focused NMR panels, enabling refined characterisation of HDL subfractions and particle distributions beyond conventional spectral measures.^20^ In addition to metabolomic features, basic clinical and demographic variables and routine haematological parameters, including white cell count, lymphocyte count, neutrophil count, and the neutrophil-to-lymphocyte ratio, were evaluated. These variables did not rank among the dominant predictors and did not materially influence model performance. The combined untargeted NMR metabolome and targeted NMR-derived parameters (*AXINON®lipoFIT®*) retained strong discriminatory capacity, replicating findings from SCAN1 (Fig. 2). The model achieved an AUC of 0.799 (95% CI 0.793-0.805) (Fig. 2A (red), SI Fig. 7A), with an accuracy of 76.7 ± 0.2%, sensitivity of 71.7 ± 0.6%, and specificity of 77.7 ± 0.2%, closely matching performance in SCAN1 (AUC 0.83).^14^ Clear separation between cancer and non-cancer cases was observed despite increased clinical heterogeneity (Fig. 2A). Top-ranked NMR variables included glutamate (p < 0.0001), histidine (p < 0.0001), and myo-inositol (p = 0.003), which were elevated in cancer, alongside reductions in lipoprotein measures, including HDL particle concentration (HDL-p; p < 0.0001), small HDL particles (SHDL-p; p < 0.0001), and HDL cholesterol (HDL-C; p < 0.0001) (Fig. 3A-F). Odds ratio analysis (SI Table S2; Fig. 2C) demonstrated consistent pathway-level differences, with higher glutamate (OR 2.08, 95% CI 1.58-2.73), myo-inositol (OR 1.77, 95% CI 1.34-2.33), and histidine (OR 2.44, 95% CI 1.56-3.82), and lower HDL-p concentrations (OR 0.44, 95% CI 0.33-0.58) in cancer.

**Figure 2.**
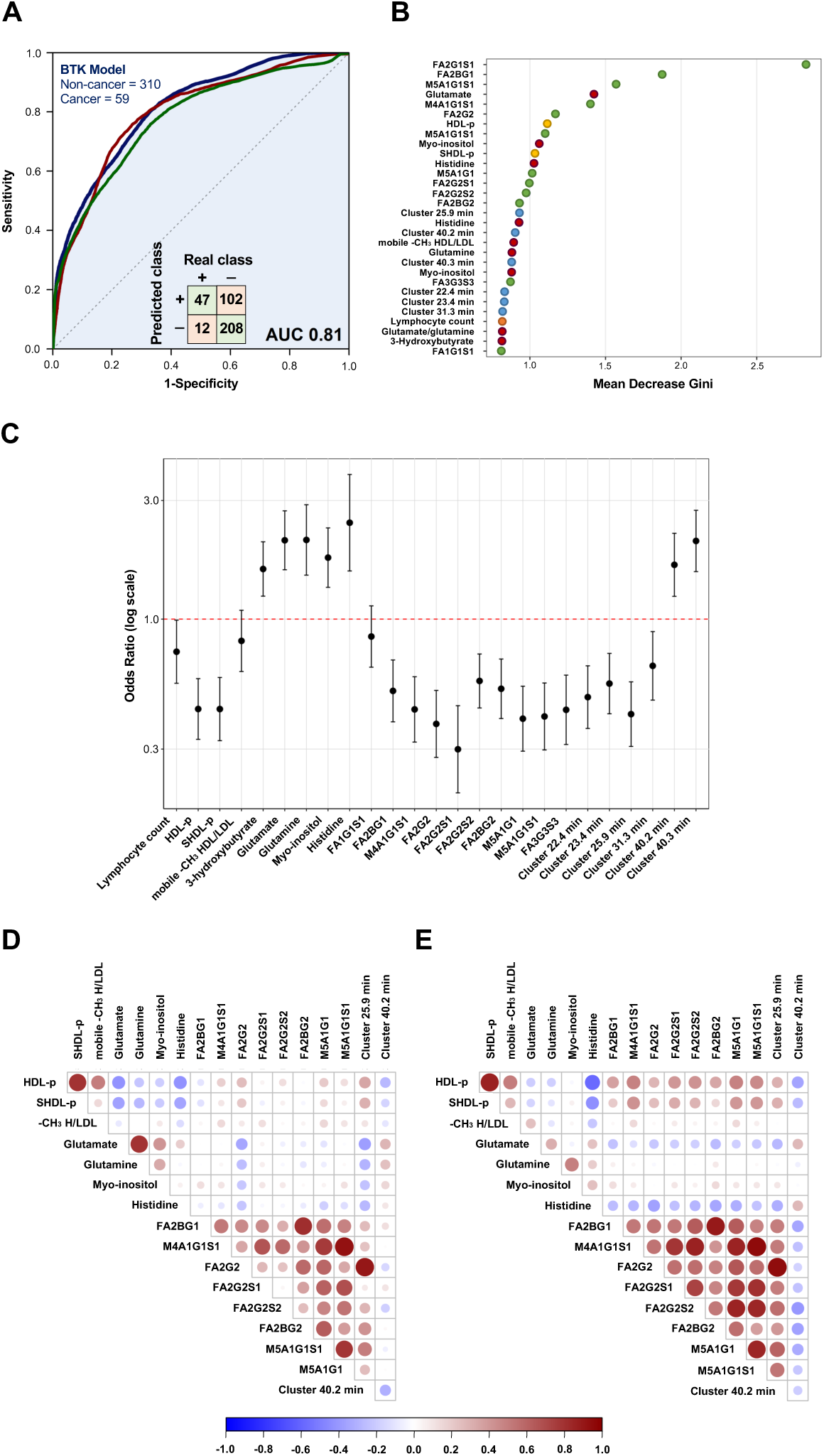
Integrated multimodal model for distinguishing cancer from non-cancer in the SCAN2 cohort. **(A)** Receiver-operating characteristic (ROC) curve for the integrated model (blue) combining untargeted NMR, targeted *AXINON®lipoFIT®*, and semi-targeted glycomics data (n = 369; 59 cancer, 310 non-cancer). Models based on NMR data alone (red) and glycomics data alone (green) are shown for comparison. The inset shows the confusion matrix at the decision threshold. **(B)** Variable-importance plot (mean decrease Gini) showing the top discriminatory features contributing to model classification. **(C)** Odds ratios (log scale ± 95% CI) for the principal variables (excluding duplicate spectral variables). **(D-E)** Correlation matrices showing pairwise relationships among the top discriminatory metabolites and glycans (excluding duplicate spectral variables). Positive correlations are depicted in red and negative correlations in blue; circle size and colour intensity indicate the strength of correlation. **(D)** Non-cancer group. **(E)** Cancer group. Metabolites and glycans formed distinct clusters, with cancer samples showing stronger inverse correlations between fucosylated glycans and HDL measures.

**Figure 3.**
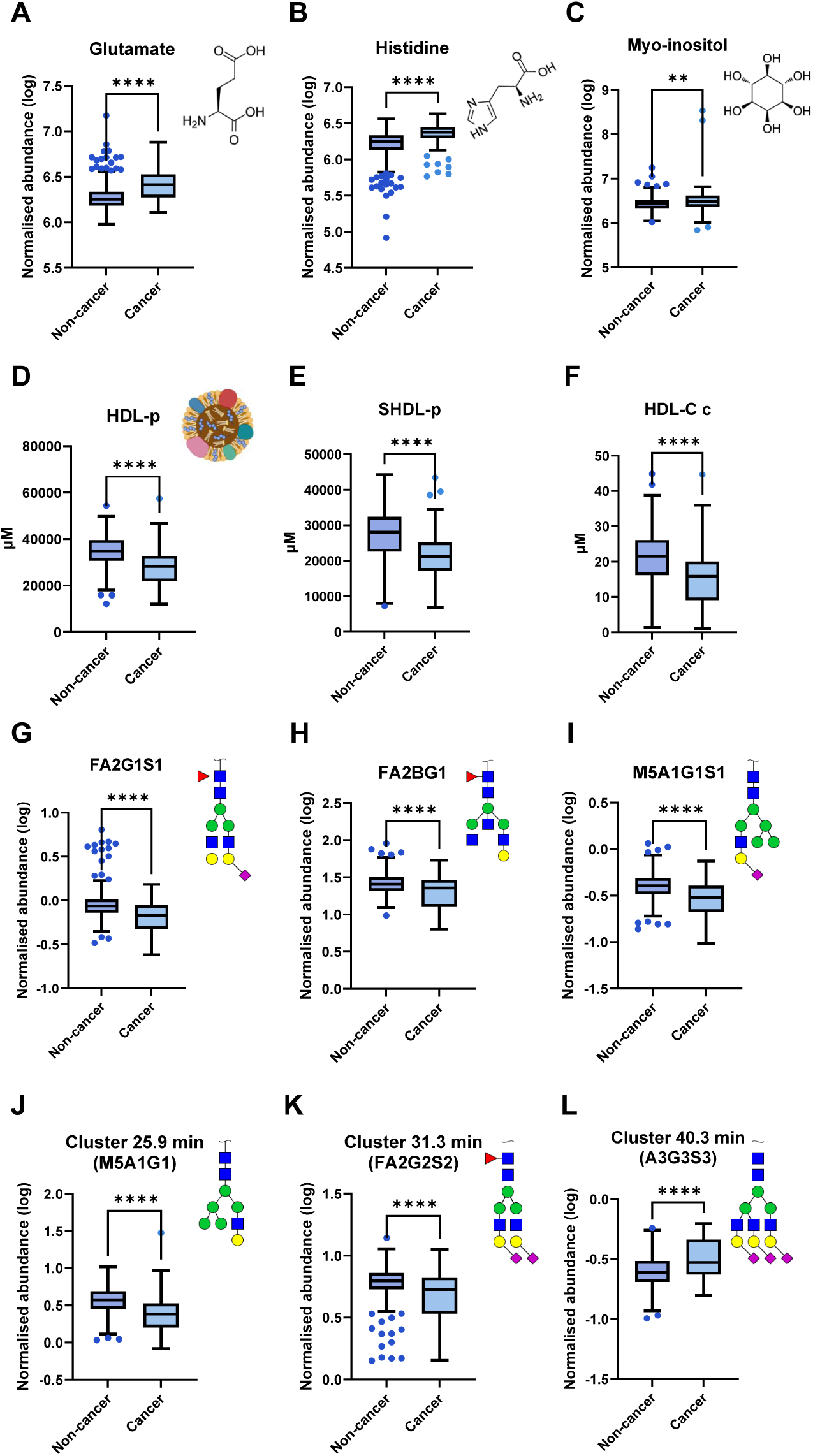
Metabolites and glycans demonstrate differential abundance between cancer and non-cancer cases. (n = 369; 59 cancer, 310 non-cancer). Boxplots show metabolites and glycans that differed significantly between cancer and non-cancer groups. (**A–F**) NMR-derived metabolites and lipoprotein measures were identified as top variables based on Gini’s index from the model including untargeted NMR bins and targeted *AXINON®lipoFIT®* parameters. (**G–I**) Glycans represent the top-ranked features from the glycomics-only model integrating fluorescence and MS data. (**J–L**) Fluorescence-derived glycan clusters correspond to retention-time peaks, with the predominant glycan structure shown for each cluster. Asterisks indicate significance (**p < 0.01; ****p < 0.0001).

To explore complementary molecular mechanisms, semi-targeted glycomic profiling was evaluated. Glycan features alone provided moderate discrimination between cancer and non-cancer cases, with a ROC AUC of 0.778 (95% CI 0.771-0.785). At the decision threshold (0.141), the model achieved a sensitivity of 76.0 ± 0.6% and a specificity of 66.9 ± 0.3%, corresponding to an overall accuracy of 68.3 ± 0.2% (Fig. 2A, green; SI Fig. 7B). Although less discriminatory than the NMR-derived metabolomic profile, glycomic features captured biologically informative alterations associated with malignancy. The most discriminatory glycans included FA2G2S1 (OR 0.299, 95% CI 0.200-0.448), FA2BG1 (OR 0.514, 95% CI 0.386-0.685), and M5A1G1S1 (OR 0.406, 95% CI 0.298-0.553), all fucosylated or bisected biantennary structures reduced in cancer (p < 0.0001; Fig. 3G-I). Consistent decreases were also observed for fluorescence-detected peaks corresponding predominantly to M5A1G1 (OR 0.415, 95% CI 0.307-0.559) and FA2G2S2 (OR 0.649, 95% CI 0.472-0.891) (both p < 0.0001; Fig. 3J). In contrast, higher-order glycans, notably A3G3S3, were increased in cancer (OR 2.062, 95% CI 1.552-2.739; p < 0.0001; Fig. 3L). The direction and magnitude of glycomic changes were coherent across platforms and inversely related to NMR-derived metabolic alterations, with reduced fucosylated and bisected structures paralleling higher glutamate, histidine, and myo-inositol, and lower HDL-p and SHDL-p concentrations. Integration of untargeted NMR metabolomics, targeted *AXINON®lipoFIT®* measurements, and semi-targeted glycomic data derived from fluorescence and mass spectrometry further improved overall model performance. The combined model achieved a ROC AUC of 0.814 (95% CI 0.808-0.820; Fig. 2A, Table 2). At the decision threshold (0.129), sensitivity reached 80.3 ± 0.5% and specificity 67.0 ± 0.3%, corresponding to an overall accuracy of 70.0 ± 0.2%. Examination of the associated confusion matrix (47 TP, 102 FP, 208 TN, and 12 FN) indicates that the operating point represents a balanced compromise between maximising cancer detection and limiting false classifications. The integrated model retained influential features from all modalities (Fig. 2B), with fucosylated and bisected glycans (FA2G2S1 (p < 0.0001, Fig. 3G), FA2BG1 (p < 0.0001, Fig. 3H), M5A1G1S1 (p < 0.0001, Fig. 3I)) ranking alongside NMR-derived metabolites (glutamate (p < 0.0001, Fig. 3A), histidine (p < 0.0001, Fig. 3B), myo-inositol (p = 0.003, Fig. 3C)) and HDL subclasses (HDL-p, SHDL-p, HDL-C (all p < 0.0001, Fig. 3D-F)). In addition, fluorescence- and MS-derived glycan clusters capturing related structural motifs (M5A1G1, FA2G2S2, and A3G3S3; Fig. 3J-L) were retained, indicating that both individual glycan species and higher-order glycan patterns contribute to model discrimination. Odds ratio analysis confirmed opposing trends across modalities, with higher cancer risk associated with elevated amino acid metabolites and lower risk linked to reduced HDL measures and fucosylated glycans. Although glycomics alone showed lower discriminatory performance, its inclusion added complementary information, reflecting coordinated metabolic and glycomic dysregulation.

**Table 2.**
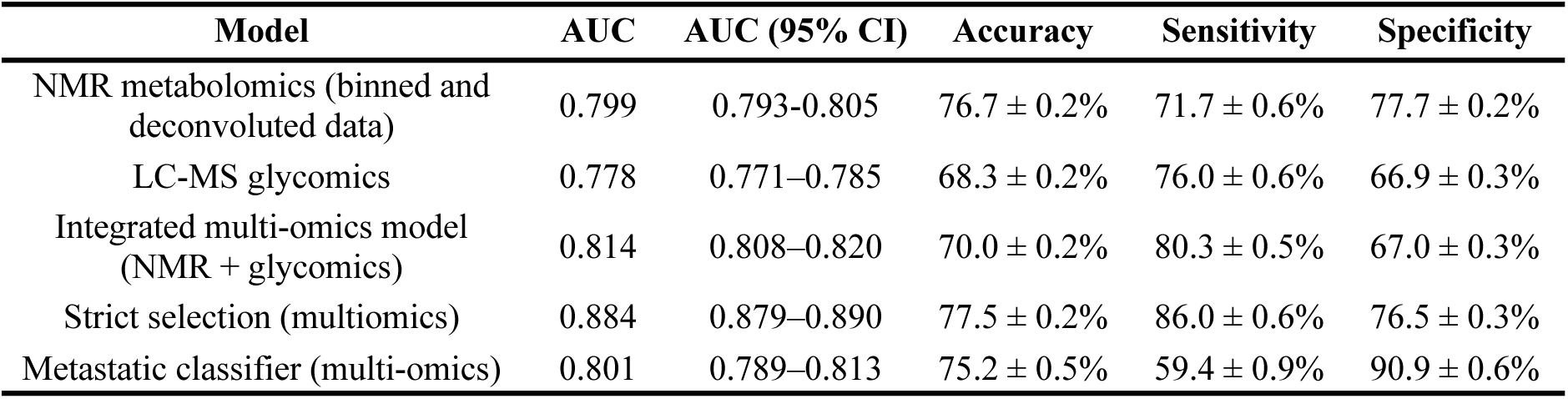
Diagnostic performance of metabolomic, glycomic, and integrated models in SCAN2.

| Model | AUC | AUC (95% CI) | Accuracy | Sensitivity | Specificity |
| --- | --- | --- | --- | --- | --- |
| NMR metabolomics (binned and deconvoluted data) | 0.799 | 0.793-0.805 | $76.7 \pm 0.2\%$ | $71.7 \pm 0.6\%$ | $77.7 \pm 0.2\%$ |
| LC-MS glycomics | 0.778 | 0.771-0.785 | $68.3 \pm 0.2\%$ | $76.0 \pm 0.6\%$ | $66.9 \pm 0.3\%$ |
| Integrated multi-omics model (NMR + glycomics) | 0.814 | 0.808-0.820 | $70.0 \pm 0.2\%$ | $80.3 \pm 0.5\%$ | $67.0 \pm 0.3\%$ |
| Strict selection (multiomics) | 0.884 | 0.879-0.890 | $77.5 \pm 0.2\%$ | $86.0 \pm 0.6\%$ | $76.5 \pm 0.3\%$ |
| Metastatic classifier (multi-omics) | 0.801 | 0.789-0.813 | $75.2 \pm 0.5\%$ | $59.4 \pm 0.9\%$ | $90.9 \pm 0.6\%$ |

Finally, to characterise relationships between individual metabolites and glycans, correlation analyses were performed separately in non-cancer and cancer groups (Fig. 2D, E). In both groups, NMR-derived metabolites (upper left quadrant) formed a distinct cluster from glycan features (lower right quadrant), indicating minimal overlap between these biochemical domains. In non-cancer samples (Fig. 2D), positive correlations were observed primarily within each modality, including among amino acids (glutamate, histidine, myo-inositol) and among structurally related glycans (FA2G2S1, FA2BG1, M5A1G1S1). In contrast, cancer samples (Fig. 2E) showed a reorganisation of correlation structure, characterised by stronger inverse associations between histidine and fucosylated and bisected glycans, alongside pronounced positive correlations between glycan signals and HDL-related measures (HDL-p, SHDL-p, and mobile lipoprotein -CH₃ signals). This pattern reflects a coordinated shift in metabolic connectivity, with altered coupling between lipid and glycan metabolism emerging specifically in malignancy. Together, these findings demonstrate that the NMR metabolome provides a reproducible diagnostic signal, with glycomic alterations contributing complementary discriminatory information. Importantly, this disease-associated signal remained robust despite substantial clinical heterogeneity, supporting its potential applicability in diverse real-world settings and providing a foundation for subsequent refined and stage-specific analyses.

### Enhanced discrimination after exclusion of major non-malignant metabolic confounders

To minimise metabolic confounding from non-malignant conditions, we evaluated a refined subgroup model (277 non-cancer; 32 cancer). In this cohort, the BTK model demonstrated significantly enhanced discriminatory performance (p < 0.0001) compared with the full-cohort model, achieving an AUC of 0.884 (95% CI 0.879-0.890), accuracy 77.5 ± 0.2%, sensitivity 86.0 ± 0.6%, and specificity 76.5 ± 0.3% (Fig. 4A, Table 2). The corresponding confusion matrix (TP = 28, FP = 65, TN = 212, FN = 4) reflects improved case detection and overall classification fidelity. Variable-importance rankings (Fig. 4B) were broadly concordant with the full cohort, with amino-acid metabolites (glutamate, histidine, myo-inositol, lactate) and HDL-related measures remaining dominant contributors, alongside several fucosylated and bisected glycans (SI Table 3).

**Figure 4.**
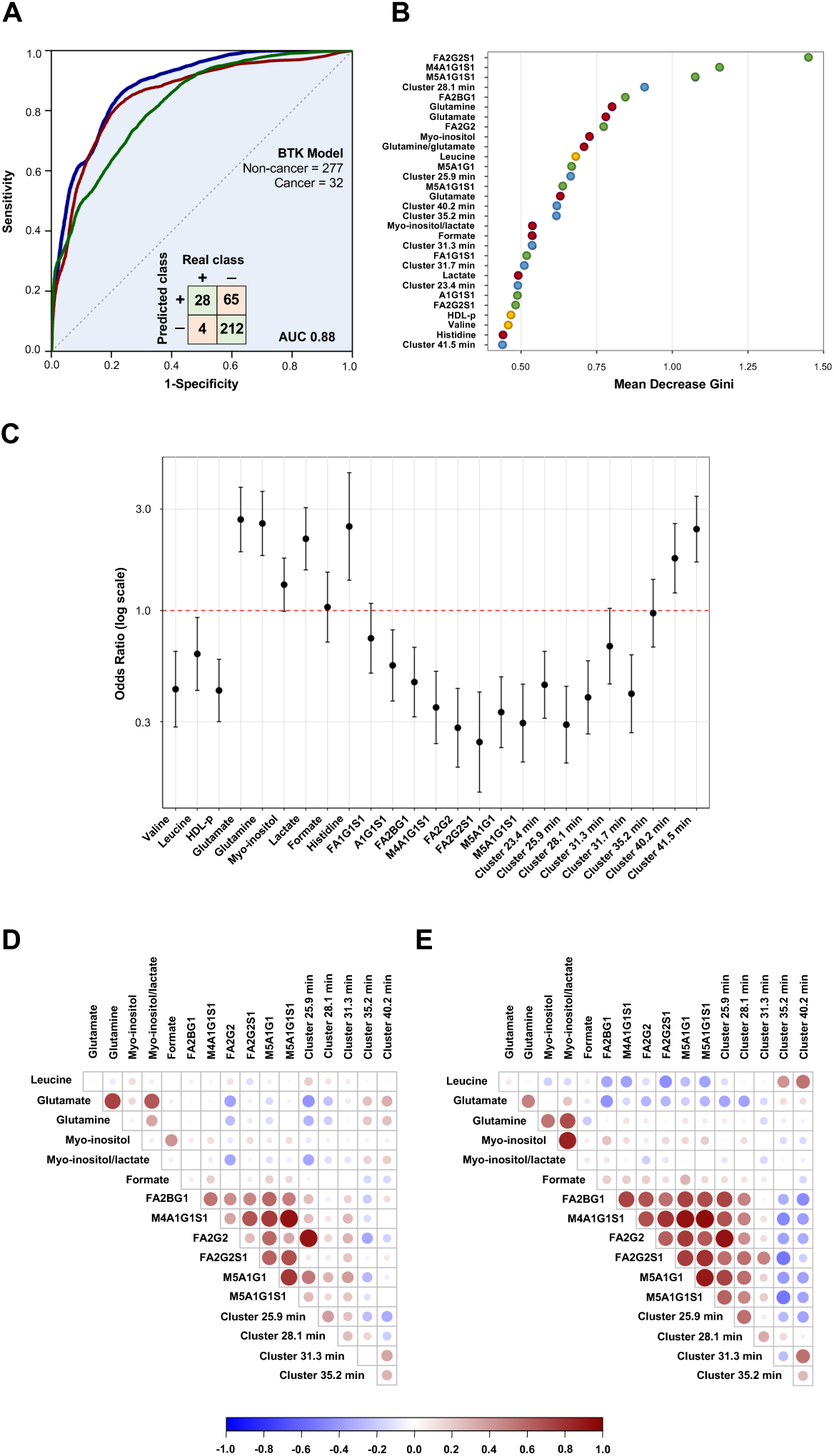
Integrated multimodal model for distinguishing cancer from non-cancer in the more strictly selected subset of the SCAN2 cohort (for exclusion criteria see. Fig. 1**). (A)** Receiver-operating characteristic (ROC) curve for the integrated model (blue) combining untargeted NMR, targeted *AXINON®lipoFIT®*, and semi-targeted glycomics data (n = 309; 32 cancer, 277 non-cancer). Models based on NMR data alone (red) and glycomics data alone (green) are shown for comparison. The inset shows the confusion matrix at the decision threshold. **(B)** Variable-importance plot (mean decrease Gini) showing the top discriminatory features contributing to model classification. **(C)** Odds ratios (log scale ± 95% CI) for the principal variables (excluding duplicate spectral variables). **(D-E)** Correlation matrices showing pairwise relationships among the top discriminatory metabolites and glycans (excluding duplicate spectral variables). Positive correlations are depicted in red and negative correlations in blue; circle size and colour intensity indicate the strength of correlation. **(D)** Non-cancer group. **(E)** Cancer group.

Odds-ratio estimates (Fig. 4C) recapitulated the effect patterns observed in the main analysis, with elevated amino-acid metabolites associated with increased odds of malignancy and reduced fucosylated glycans associated with lower odds-ratios. Correlation structures in non-cancer and cancer subgroups (Fig. 4D-E) were consistent with the full-cohort findings, demonstrating preserved modality-specific clustering and the characteristic inverse associations between glycan features and HDL measures in cancer. Collectively, these results indicate that exclusion of major comorbidities strengthens model performance while retaining the underlying metabolic and glycomic signature of malignancy.

### Preservation of cancer probability structure across heterogeneous comorbidities

To further characterise the relationship between cancer-associated metabolic signatures and overall non-specific disease burden, we examined joint diagnosis prediction regions for cancer and alternative non-cancer diagnoses within a shared molecular feature space. Cancer probabilities were derived from the SCAN cancer model trained on the cancer sub-cohort, while non-specific disease probabilities were generated using an independent classifier trained on alternative non-cancer diagnosis status, but constrained to the same metabolomic variables.

Projection of individuals into the joint logit-transformed probability space showed clear separation between cancer and non-cancer participants along the cancer probability axis (Fig. 5A, B), consistent with the strong discriminatory performance of the cancer model. Cancer cases clustered within a relatively compact region of the joint space, as reflected by a well-defined 75% kernel density contour. In contrast, predictions for alternative non-cancer diagnoses showed substantially greater dispersion and overlap (Fig. 5C, D), consistent with the biological and clinical heterogeneity of the non-malignant conditions represented in the cohort, including cardiovascular, renal, and respiratory disease. Despite this heterogeneity, the cancer-associated metabolic signal remained stable across the full spectrum of predicted comorbidity risk, indicating that cancer-related metabolic perturbations were preserved even in multimorbid individuals.

**Figure 5.**
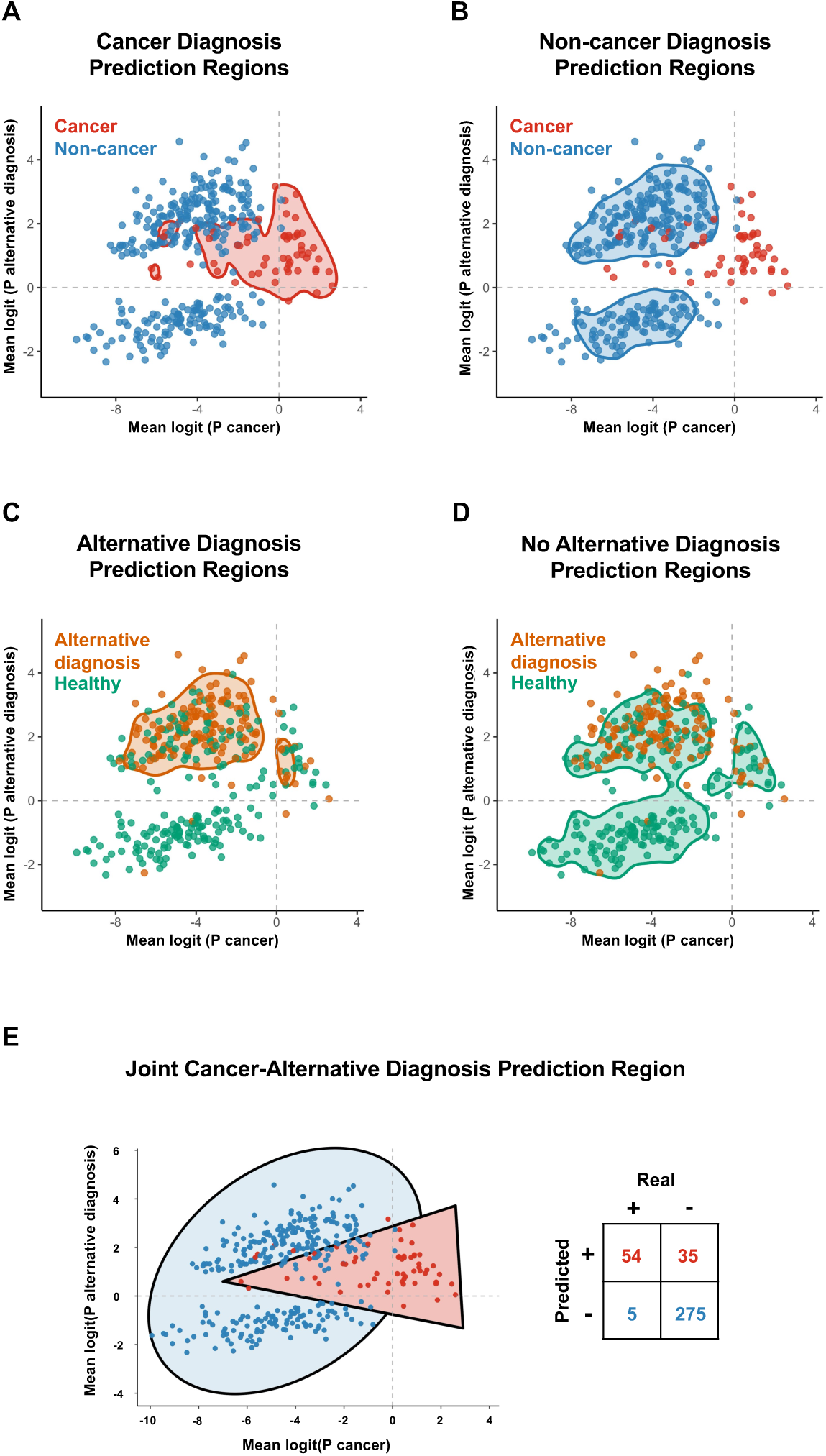
Joint probability representation of cancer and alternative non-cancer diagnosis risk in the SCAN2 cohort. Participants are mapped into a joint logit-transformed probability space defined by predicted cancer risk and alternative non-cancer diagnosis risk. Predictions were generated by independent classifiers trained on a shared metabolomic feature set. Points are coloured by observed cancer status **(A, B)** or alternative non-cancer diagnosis **(C, D)**. Kernel density-based contours enclose 75% of the joint probability mass, summarising dominant patterns of sample concentration without reliance on fixed probability cut-offs. Dashed lines denote zero on the logit scale. In the joint space **(E)**, a directional wedge-shaped region illustrates a projection-based decision rule aligned with increasing cancer probability while accounting for comorbidity burden. This region was used to derive the confusion matrix and summary metrics, providing an interpretable illustration of classification behaviour within the joint probability space rather than an independent validation of diagnostic performance.

To enable downstream classification and quantitative assessment, an explicit geometric decision region was defined within the joint probability space. A directional wedge-shaped region was constructed to capture the dominant axis of increasing cancer probability while accounting for concurrent alternative non-cancer diagnostic burden (Fig. 5E). Individuals within this region were classified as predicted cancer-positive, allowing construction of a confusion matrix and calculation of performance metrics. Under this projection-based decision rule, high overall accuracy was observed (88.9%), with balanced sensitivity (89.8%) and specificity (88.7%) (Fig. 5E). These metrics reflect the internal consistency of cancer probability estimates within the joint space and illustrate how the continuous risk landscape can be mapped onto a discrete classification framework. Importantly, this analysis provides an interpretable representation of joint cancer-alternative non-cancer diagnosis risk rather than an independent validation of diagnostic performance; all probability estimates were derived from cross-validated predictions with strict separation of training and test data throughout.

### High-specificity discrimination of metastatic disease in a non-specific symptom cohort

Integrated metabolomic and glycomic profiling enabled discrimination between metastatic and non-metastatic cancer. The multivariable classification model achieved a ROC AUC of 0.801 (95% CI 0.789-0.813), with an overall accuracy of 75.2 ± 0.5% (Fig. 6A, Table 2). At the decision threshold (0.575), the BTK model showed high specificity for non-metastatic disease (90.9 ± 0.6%) and more moderate sensitivity for metastatic disease (59.4 ± 0.9%). In absolute terms, this corresponded to 27 TN and 17 TP, with 3 FP and 12 FN. This performance profile reflects a clinically relevant trade-off in a non-specific symptom population, prioritising reliable exclusion of advanced malignancy while retaining meaningful sensitivity for metastatic disease.

**Figure 6.**
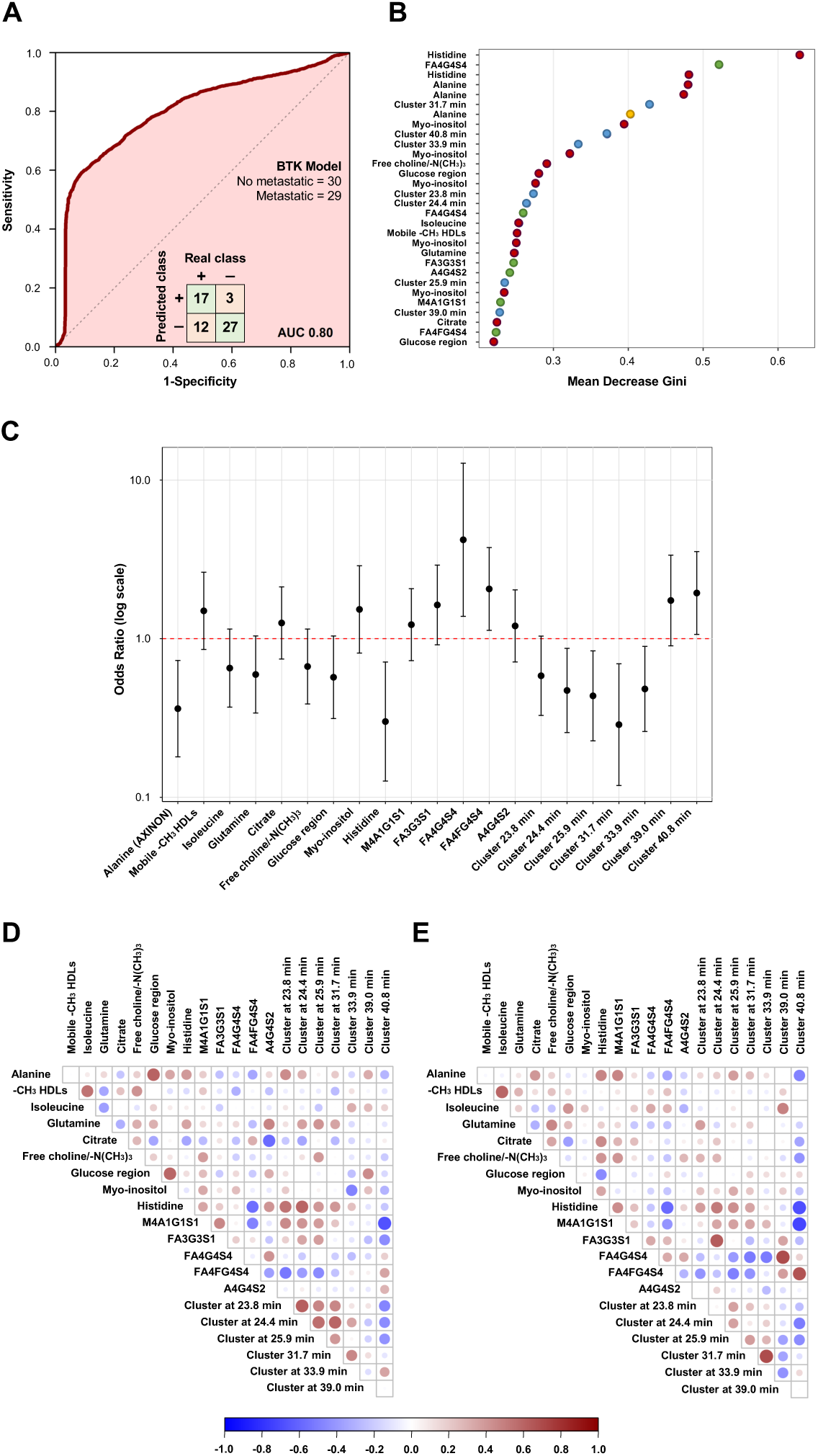
Integrated multimodal model for distinguishing metastatic from non-metastatic cancer in the SCAN2 cohort. **(A)** Receiver-operating characteristic (ROC) curve for the integrated classification model combining untargeted NMR metabolomics, targeted *AXINON®lipoFIT®* metabolite quantification, and semi-targeted *N*-glycan profiling. The inset shows the confusion matrix at the decision threshold. **(B)** Variable-importance plot based on mean decrease in Gini index, showing the top features contributing to model classification, including amino acids, lipoprotein-derived NMR signals, spectral regions, and glycan-associated features. **(C)** Univariable odds ratios (log scale, points) with 95% confidence intervals (error bars) for the principal discriminatory variables (excluding duplicate spectral representations), indicating the direction and strength of association with metastatic disease. **(D-E)** Pairwise correlation matrices of the top discriminatory features (excluding duplicate spectral variables) stratified by disease status. Positive correlations are shown in red and negative correlations in blue, with circle size and colour intensity proportional to correlation strength. **(D)** Non-metastatic cancer group. **(E)** Metastatic cancer group.

Variable importance analysis from the integrated random forest model identified a heterogeneous panel of discriminatory features spanning amino acids, lipoprotein-derived NMR signals, spectral regions, and *N*-glycan measures (Fig. 6B; SI Table 4). The highest-ranked variables by mean decrease in Gini index included histidine, the complex tetra-antennary glycan FA4G4S4, alanine, and a fluorescence-derived glycan cluster eluting at 31.7 min, indicating contributions from both small-molecule metabolism and higher-order glycan structures. Additional features with stable multivariate importance comprised myo-inositol, mobile -CH₃ HDL signals, glucose and free choline spectral regions, and multiple glycan-associated chromatographic clusters, consistent with the distributed nature of the metastatic signature.

Univariable analysis confirmed that several of the top-ranked features were individually associated with metastatic status (Fig. 6C, 7). Histidine showed a significant inverse association with metastatic disease (OR 0.41, 95% CI 0.21-0.81; p = 0.018; Fig. 7A), as did alanine measured by both untargeted NMR and targeted *AXINON®lipoFIT®* quantification (OR 0.36, 95% CI 0.18-0.73; p = 0.015; Fig. 7B, D). Myo-inositol, SHDL-p, and SLDL-p were also ranked among the most discriminatory features and showed weaker associations with metastatic cancer that did not survive multiple-testing correction (Fig. 7C, E, F). In contrast, selected *N*-glycan features were positively associated with metastatic status, including FA4G4S4 quantified by fluorescence detection (OR 2.33, 95% CI 1.25-4.33; p = 0.018) and by mass spectrometry (OR 4.20, 95% CI 1.38-12.76; adjusted p = 0.024). Fluorescence-derived glycan clusters eluting at 31.7 min and 40.8 min also remained significant following adjustment (p = 0.015 and 0.050, respectively). Although several additional metabolites, lipoprotein measures, and glycan clusters did not retain statistical significance after multiple-testing correction, their effect directions were concordant with multivariate importance rankings. Together, these findings indicate that discrimination between metastatic and non-metastatic disease arises from coordinated alterations across amino acid metabolism, lipoprotein-associated signals, and complex *N*-glycan architecture, supporting the value of integrative multi-omic modelling for stratifying disease burden.

**Figure 7.**
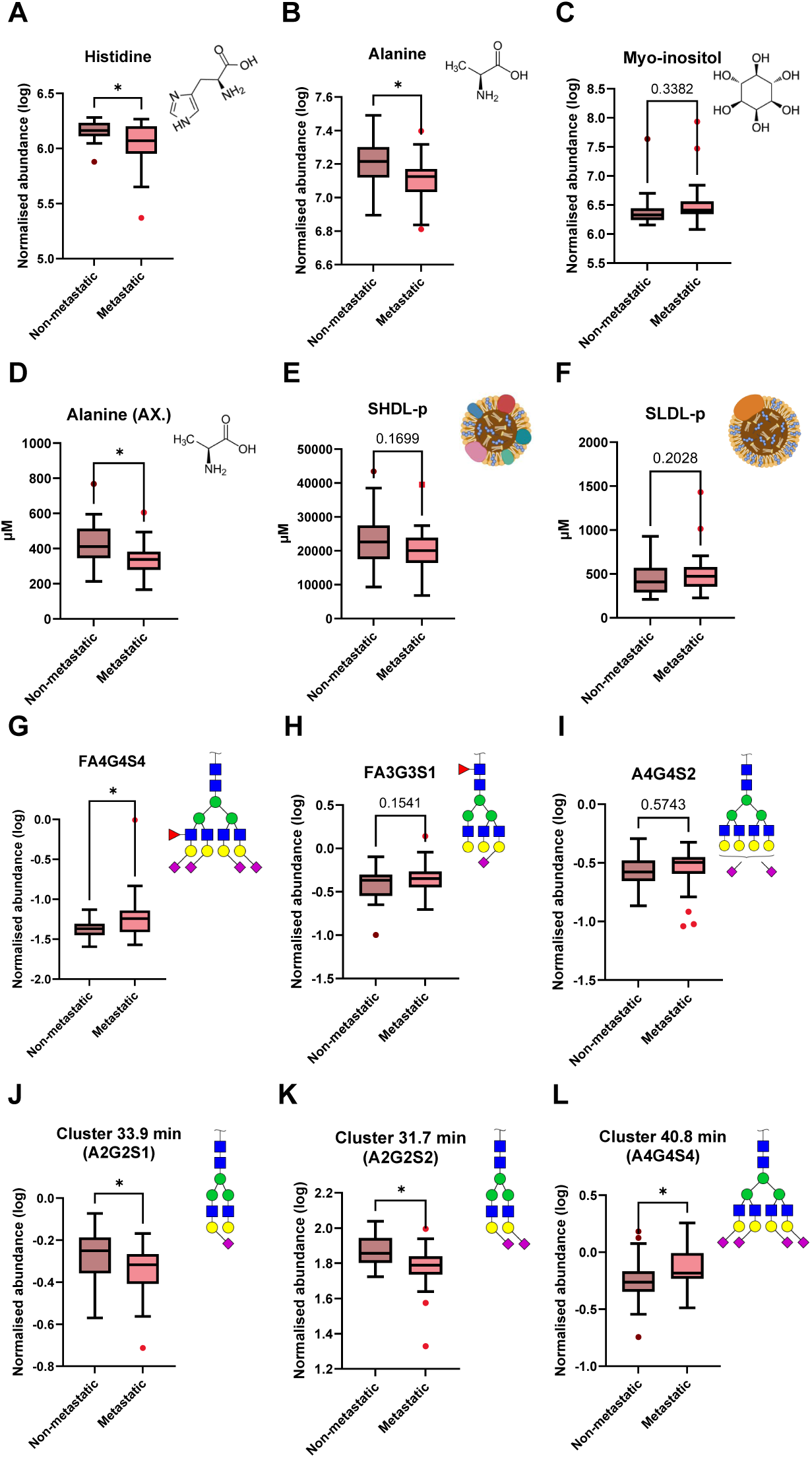
Metabolites and glycans demonstrate differential abundance between metastatic and non-metastatic cancer. (n = 59; 29 metastatic cancer, 30 non-metastatic cancer). Boxplots show metabolites and glycans that differed significantly between metastatic and non-metastatic disease. **(A-F)** NMR-derived metabolites and lipoprotein measures were identified as top variables based on Gini’s index from a model integrating untargeted NMR spectral bins with targeted *AXINON®lipoFIT®* parameters, with small HDL particle concentration (SHDL-p) and small LDL particle concentration (SLDL-p) emerging as the top *AXINON®lipoFIT®*-derived features. **(G-I)** Glycans represent the top-ranked features from the glycomics-only model integrating fluorescence and mass spectrometry data. **(J-L)** Fluorescence-derived glycan clusters correspond to retention-time peaks, with the predominant glycan structure shown for each cluster. Asterisks indicate statistical significance (*p < 0.05; **p < 0.01).

Correlation analyses revealed marked differences in feature inter-relationships between non-metastatic and metastatic groups (Fig. 6D-E). Non-metastatic cases showed relatively structured correlation patterns, with moderate positive associations among amino acids and lipid-associated NMR signals (Spearman |r| ≈ 0.3-0.6). In contrast, metastatic disease was characterised by a reorganisation of correlation structure, including attenuation of metabolite-lipoprotein associations and strengthening of glycan-glycan correlations, several exceeding |r| ≈ 0.5, consistent with coordinated metabolic and glycomic reprogramming. Collectively, these coordinated alterations underpin the multivariate metastatic signature, yielding high specificity for non-metastatic disease (90.9%) and moderate sensitivity for metastatic disease (59.4%), supporting the utility of integrated metabolomic-glycomic profiling for disease stratification in non-specific symptom populations.

## DISCUSSION

We demonstrate that malignancy is associated with a distinct and reproducible systemic molecular phenotype that can be distinguished from non-specific illness in patients presenting through real-world diagnostic pathways.^31^ Evidence for this phenotype was observed consistently across independent cohorts analysed using different platforms and modelling strategies. Such replication is notable given the limited clinical translation of many previously reported cancer metabolomic signatures, which has often been attributed to insufficient external validation, biological ambiguity, and analytical variability.^32^ Consistent with critiques of NMR-based cancer metabolomics, which emphasise that circulating metabolites reflect whole-organism physiology rather than tumour metabolism alone,^32,33^ our findings support a conserved host metabolic response to malignancy. A cancer-associated metabolic signature originally reported in the published SCAN1 cohort^14^ was independently reproduced in the larger and clinically more heterogeneous SCAN2 cohort, despite the use of a different analytical pipeline and pre-analytical controls. Conversely, metabolites independently prioritised in SCAN2 retained discriminatory performance and concordant effect directions when re-evaluated in SCAN1. Importantly, SCAN2 data were acquired on a commercial FDA-cleared 600 MHz *AXINON®lipoFIT®* NMR platform,^20^ arguing against cohort-, platform-, or procedure-specific effects. Together, this reciprocal replication across cohorts, platforms, and analytical strategies provides strong evidence that the observed metabolic phenotype reflects a robust and generalisable systemic response to malignancy, rather than artefacts of study design or analytical workflow.

A key advance of SCAN2 is the integration of orthogonal glycomic profiling alongside metabolomics. Aberrant glycosylation is a recognised hallmark of malignancy, with systemic consequences reflected in circulating acute-phase proteins, immunoglobulins, and hepatic secretory glycoproteins.^34,35^ These changes are only indirectly captured by conventional NMR metabolomics, which primarily resolves small molecules and lipid subfractions. Incorporation of LC-MS-based glycan analysis therefore interrogates a complementary biochemical axis,^36^ yielding discriminatory features that are additive rather than redundant. The improved performance of the integrated model demonstrates that malignancy perturbs multiple molecular systems detectable at first clinical presentation, supporting the value of a multi-omics approach in non-site-specific diagnostic settings.

Closer examination of the discriminatory features provides insight into the biological basis of model performance. The enrichment of complex biantennary *N*-glycans with core fucosylation and terminal sialylation (FA2G2S1, FA2G2S2, FA2BG1, and FA2BG2) is consistent with cancer-associated remodelling of circulating glycoproteins driven by inflammatory cytokine signalling and hepatic acute-phase responses.^37–40^ Such increases in sialylation and fucosylation have been reported across multiple malignancies and are generally interpreted as markers of host immune modulation rather than tumour-derived secretion.^37,38,41^ The presence of bisecting GlcNAc-containing structures further supports immune remodelling, consistent with altered immunoglobulin function and inflammatory signalling in cancer.^40,42^

Detection of high-mannose and hybrid-type glycans (M5A1G1, M4A1G1S1) indicates disrupted glycan processing and increased protein turnover, features associated with systemic inflammation and malignancy.^43–45^ In blood, these structures are typically interpreted as markers of dysregulated glycoprotein biosynthesis rather than direct tumour products, and their coexistence with fully sialylated glycans suggests a globally perturbed glycosylation landscape.^44^ These glycomic changes converge with NMR-derived markers of glycoprotein acetylation and lipoprotein remodelling.^46,47^ GlycA and GlycB index composite *N*-acetyl signals from circulating acute-phase proteins (α1-acid glycoprotein, haptoglobin, and α1-antitrypsin) and are linked to cancer and chronic inflammatory states,^48^ while concurrent alterations in HDL particle profiles are consistent with inflammation-driven changes in hepatic lipid handling.^47^

The accompanying metabolic perturbations, including altered histidine, glutamate, and myo-inositol levels, were consistently observed across both SCAN1 and SCAN2, supporting their interpretation as components of a systemic host response to malignancy.^49,50^ Reduced circulating histidine has been widely reported in cancer and inflammatory states and is thought to reflect increased utilisation during immune activation and oxidative stress.^50,51^ Dysregulated glutamate is compatible with altered amino-acid flux and energy metabolism characteristic of cancer-associated metabolic reprogramming, while changes in myo-inositol have been linked to hepatic dysfunction, insulin signalling, and inflammation.^52^ Together, the coordinated disruption of glycomic, lipoprotein, and metabolic features points to convergent perturbation of inflammatory, hepatic, and metabolic pathways,^46^ providing a biologically plausible basis for the robustness of the integrated model across heterogeneous clinical presentations.

A distinctive contribution of this study is the use of orthogonal biplot representations to separate molecular signals associated with general illness from those specific to malignancy. Rather than relying on a single cancer-versus-all classifier, the modelling framework explicitly disentangles non-specific illness and cancer-associated perturbation by training independent axes before projecting all individuals into a shared joint space.^30,53^ This approach reflects the clinical reality of non-site-specific referral pathways, in which many patients are systemically unwell without malignancy, and avoids conflation of illness-related and cancer-specific biology.^54^ The resulting biplots define resolvable regions in which cancer cases cluster independently of the severity of generic illness, indicating that discrimination is driven by malignancy-associated metabolic and glycomic features rather than by overall systemic disturbance.^29^ Importantly, the observed classification performance arises not simply from model complexity, but from explicit structuring of biological variance into interpretable dimensions corresponding to illness and cancer.^29^ This representation enhances transparency and clinical plausibility by allowing individual patients to be contextualised across multiple biological processes rather than reduced to a single composite score.

The ability of the integrated approach to discriminate metastatic from non-metastatic disease represents an important secondary finding. Although sensitivity was lower than specificity, the strong rule-out performance is clinically relevant. As well as being highly reassuring for cancer survivors, patients presenting with non-specific symptoms often undergo extensive imaging to exclude disseminated disease.^3^ A blood-based signature that reliably identifies patients unlikely to harbour metastatic spread could help streamline diagnostic pathways and reduce delay, particularly in resource-constrained settings.^7,54^ These observations are consistent with SCAN1 and support the interpretation that systemic metabolic disturbance scales with tumour burden.^14^

Several considerations temper these conclusions. Although this study represents a substantial expansion of the original SCAN cohort,^14^ the number of cancer cases - and particularly metastatic cases - remains modest, necessitating validation in larger independent datasets to confirm generalisability and refine model calibration.^28^

While pre-analytical handling was more rigorously standardised than in SCAN1, some real-world variability is unavoidable in fast-track diagnostic pathways; however, replication of the metabolic signature under these conditions supports biological robustness and clinical feasibility rather than undermining it. The integrated multi-omics model also incorporates a relatively large number of features across NMR and LC-MS platforms. Although this enhances biological coverage and performance,^55,56^ future work should assess whether a reduced panel of metabolites and glycans could retain comparable accuracy while facilitating translation into routine clinical practice, particularly given the current requirement for specialised glycomic infrastructure.^57^ Finally, although patients with active cancer treatment or organ-specific diagnostic pathways were excluded, residual effects of comorbidity, medication use, or diet cannot be fully excluded.^58^ Such influences are inherent to real-world cohorts and highlight the importance of prospective evaluation across diverse populations.^59^

This study demonstrates that integrated NMR metabolomics and LC-MS glycomics can improve early cancer detection in patients presenting to primary care with non-specific symptoms, a group at high risk of diagnostic delay. Replication of the SCAN1 metabolic signature in a larger, clinically heterogeneous cohort indicates a stable underlying biological signal across populations and sampling conditions. The addition of glycomic profiling enhances diagnostic performance and insight into systemic inflammatory and metabolic perturbations associated with malignancy, while orthogonal biplot representations separate cancer-specific biology from generic illness, improving interpretability in heterogeneous clinical settings. Together, these findings support blood-based multi-omics assessment as a feasible adjunct to existing diagnostic pathways, providing objective biological information to support risk stratification in real-world primary care.

## Contribution statement

D. C. A., J. R. L., and S. A. conceived and designed the study. D. C. A., S. A., and the SCAN consortium were responsible for study oversight and patient recruitment. A. G. Y. and S. A. were involved in sample collection and processing. A. G. Y. and S. de J. performed NMR experiments. P. L. H. G. prepared the samples for glycomics analysis. J. C. and G. E.-H. performed the glycomics experiments and data processing. E. S. oversaw the NMR analysis. D. I. R. S. oversaw the glycomics analysis. T.K. and J. R. L. processed and analysed the data. T. K., J. J. M., and B. S. performed the multivariate analysis. T. K. drafted the manuscript. D. C. A. provided critical feedback and guidance for analysis design and manuscript writing. All authors reviewed and approved the final manuscript.

## The SCAN Consortium

Hasan Al-Sattar, Niamh Appleby, Guanke Bao, Gagandeep Batth, Rachel Benamore, Andrew Blake, Georgeta Ciuban, Kiana K Collins, Isabella de Vere Hunt, Helene Dreau, Monica Evans, Nick Gibb, Fergus Gleeson, Shelley Hayles, Laura Heath, Ashley Jackson, Catherine Johnson, Zoe Kaveney, David Lewis, Rosie Lomas, Daniel Mitchener, Alexandra Montagu, Julia-Ann Moreland, Niall Moore, Brian Nicholson, Sophie Roberts, Neelaksh Sadhoo, Shyamal Saujani, Anna Schuh, Andrew Slater, Sarah Smith, Nia Taylor, Sharon Tonner, Suzanne Waterfield, Louise Wing.

## Ethics approval and consent to participate

Written informed consent was obtained from all participants. The study was conducted in accordance with the UK Policy Framework for Health and Social Care Research and received ethical approval from the Oxford Radcliffe Biobank research tissue bank (20/A120). This study followed the **RE**porting of studies **C**onducted using **O**bservational **R**outinely-collected **D**ata (**RECORD**) reporting guidelines.

## Data access and availability

Anonymised data not published within this article will be made available by request to the corresponding authors from any qualified investigator.

## Acknowledgement

The authors thank all patients who participated in this study.

