## Supplementary Information for "Integrating NMR Metabolomics and Glycomics for Early Cancer Detection in Patients with Non-Specific Symptoms"

Tereza Kacerova^1,2,3*, †^, Abi G. Yates^4†^, James R. Larkin^4^, Boris Shulgin^4^, Jack J. J. J. Miller^5,6^, Philippa L. Harris Gleave^4^, Sebastian de Jel^7^, Jack Cheeseman^8^, Georgia Elgood-Hunt^8^, the SCAN Consortium*^#^*, Eric Schiffer^7^, Daniel I. R. Spencer^8^, Suzie Anthony^9^, Daniel C. Anthony^4*^

*^1^ Chemistry Research Laboratory, Department of Chemistry, University of Oxford, Oxford OX1 3TA, UK*

*^2^ Kavli Institute of Nanoscience Discovery, Dorothy Crowfoot Hodgkin Building, Oxford OX1 3QU, UK*

*^3^ Physical and Theoretical Chemistry, University of Oxford, Oxford OX1 3QZ, UK*

*^4^ Department of Pharmacology, University of Oxford, Oxford OX1 3QT, UK*

*^5^ Oxford Centre for Magnetic Resonance Research, University of Oxford, Oxford OX3 9DU, UK*

*^6^ Department of Clinical Medicine, Aarhus University, Aarhus 8200, Denmark*

*^7^ numares AG, Regensburg 93053, Germany*

*^8^ Ludger Ltd, Culham Science Centre, Abingdon OX14 3EB, UK*

*^9^ Interventional Oncology, Department of Radiology, University of Oxford, Oxford OX3 9DU, UK*

** Corresponding authors*

***Daniel C. Anthony (clinical and biological)***

*Department of Pharmacology, University of Oxford, Oxford OX1 3QT, UK*

***Tereza Kacerova (analytical chemistry and data analysis)***

*Chemistry Research Laboratory, Department of Chemistry, University of Oxford, Oxford OX1 3TA, UK*

†These authors contributed equally

\# Members listed at the end of the Article

**Demographics**

SI Table 1 **Distribution of cancer diagnoses in the SCAN2 cohort.** Cancer types are shown with corresponding metastatic status at the time of assessment and participant counts. The cohort comprises a broad range of solid and haematological malignancies, reflecting the heterogeneity of a non-site-specific cancer detection setting. Metastatic status was defined according to clinical staging and imaging data available at recruitment.

| **Cancer** | **Metastatic** | **Count** |
| --- | --- | --- |
| Adenocarcinoma (not specified) | yes | 4 |
| Adenocarcinoma (colon) | yes | 2 |
| Adenocarcinoma (ovarian) | yes | 1 |
| Adenocarcinoma (lung) | no | 1 |
| Adenocarcinoma (pancreas) | yes | 1 |
| Adenocarcinoma (pancreas) | no | 2 |
| Adenocarcinoma (prostate) | yes | 2 |
| Adenocarcinoma (prostate) | no | 3 |
| B cell lymphoma | no | 1 |
| B cell lymphoma | yes | 6 |
| Basal cell carcinoma | no | 2 |
| Bladder cancer | no | 1 |
| Breast carcinoma | no | 1 |
| Carcinoid | no | 1 |
| Cervical cancer | yes | 1 |
| Cholangiocarcinoma | yes | 2 |
| Clear cell carcinoma kidney | yes | 1 |
| Ductal carcinoma in situ | no | 1 |
| Endometroid endometrial carcinoma grade 1 | no | 1 |
| Epithelioid mesothelioma | yes | 1 |
| Hepatocellular carcinoma | no | 1 |
| Gastrointestinal stromal tumour | no | 1 |
| Lung cancer | yes | 1 |
| Lung cancer | no | 1 |
| Mesothelioma | no | 1 |
| Myelodysplasia | no | 1 |
| Myeloma | no | 1 |
| Neuroendocrine | no | 1 |
| Pancreatic | yes | 1 |
| Papillary transitional cell carcinoma | yes | 1 |
| Prostate cancer | no | 6 |
| Prostate cancer | yes | 1 |
| Renal cell carcinoma | no | 1 |
| Renal cell carcinoma | yes | 3 |
| Squamous cell carcinoma | no | 1 |
| Unknown primary cancer | yes | 1 |
| Unknown cancer | no | 1 |

**Assessment of age as a potential confounder**

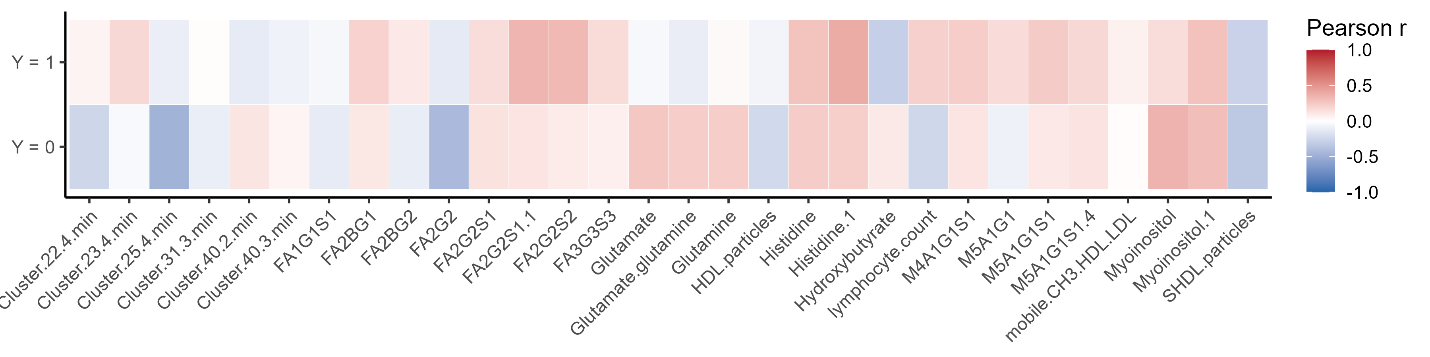

SI Figure 1 **Assessment of age as a potential confounder of model-derived features.** Heatmap showing Pearson correlation coefficients between age at SCAN and the highest-ranked metabolic and glycomic variables contributing to the cancer classification model, stratified by outcome group (Y = 1, cancer; Y = 0, non-cancer). Colour intensity reflects the direction and magnitude of correlation (red, positive; blue, negative). Across both outcome groups, correlations between age and model-informative variables were uniformly weak (|r| < 0.35), indicating that model discrimination was not driven by age-related effects.

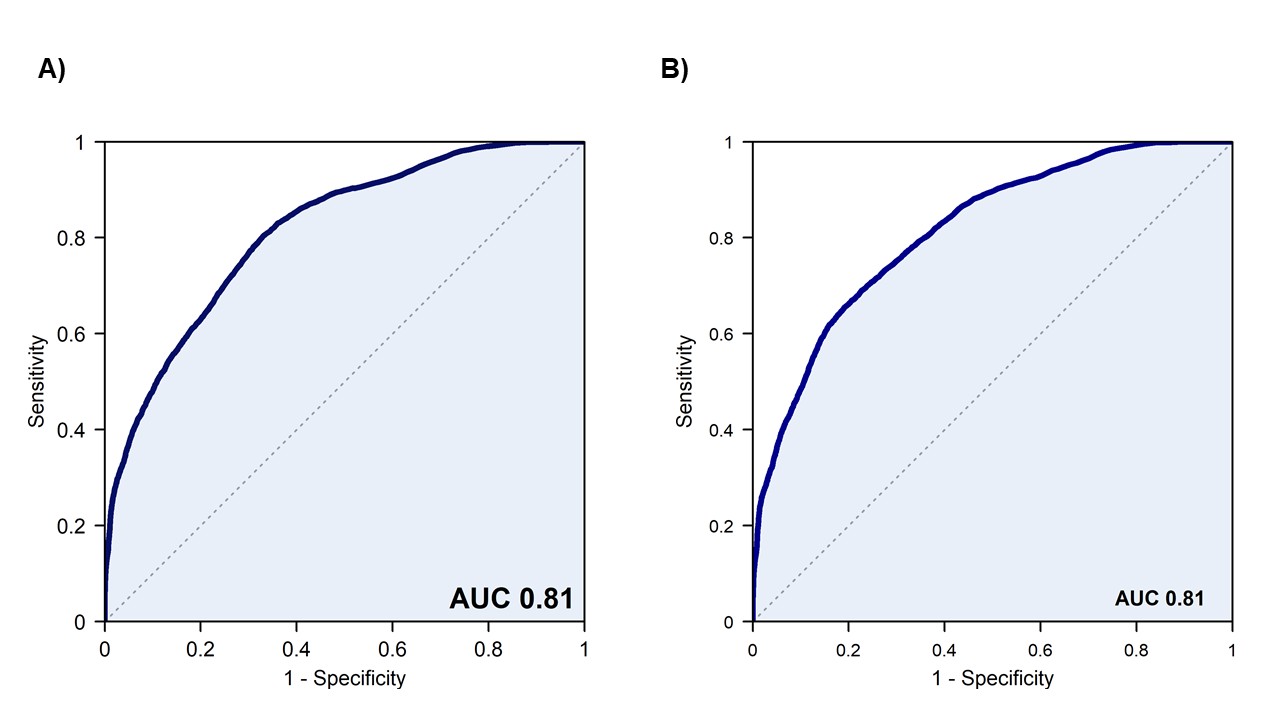

**SI Figure 2 Effect of age inclusion on model discrimination in the SCAN2 cohort.** Receiver operating characteristic (ROC) curves showing model performance when all variables, including age, are included **(A)** and when age is excluded **(B)**. Both models demonstrate identical discrimination (AUC = 0.81), indicating that inclusion of age does not materially influence overall model performance.

**Assessment of age as a potential confounder**

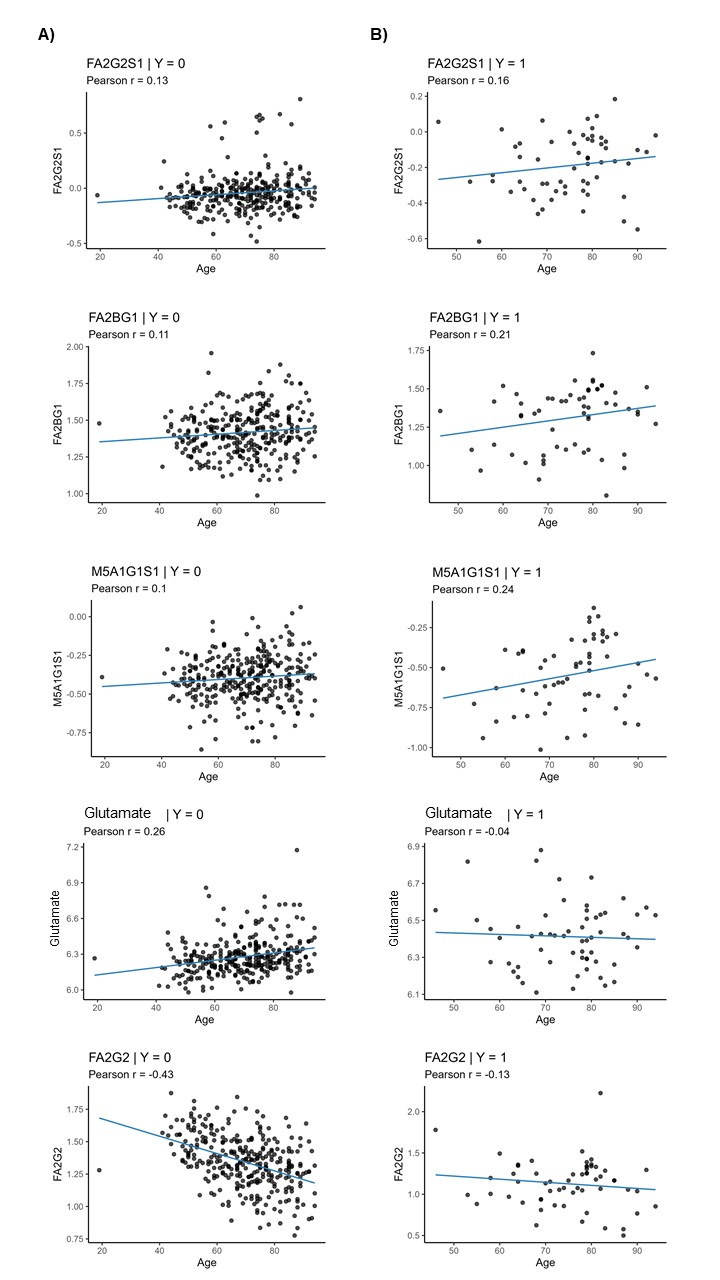

SI Figure 3 **Representative Pearson correlation analyses between age and model-informative features.** Scatter plots illustrate the relationship between age at SCAN and selected high-ranking variables from the cancer classification model, shown separately for non-cancer (Y = 0, (**A**)) and cancer (Y = 1, (**B**)) participants. Solid lines denote linear fits, with Pearson correlation coefficients indicated in each panel. Across both outcome groups, correlations were weak and inconsistent in direction, supporting minimal age dependence of individual model features.

**Assessment of age as a potential confounder**

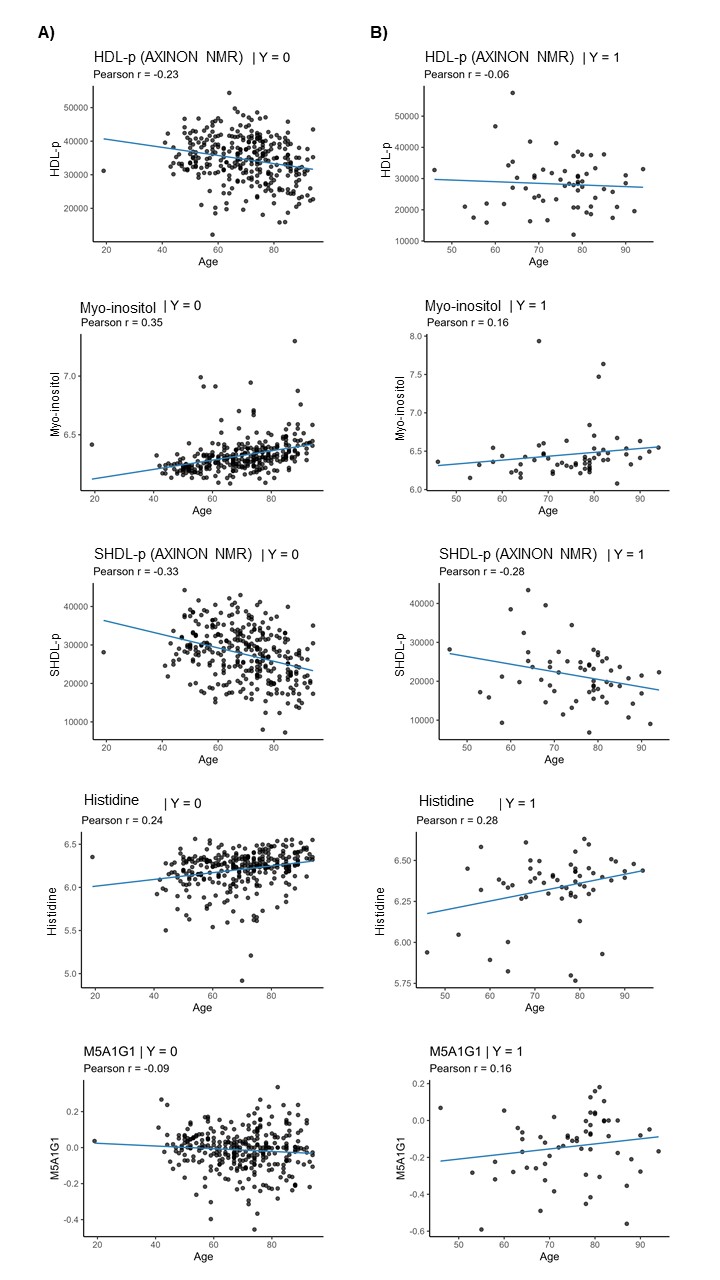

SI Figure 4 **Representative Pearson correlation analyses between age and model-informative features.** Scatter plots illustrate the relationship between age at SCAN and selected high-ranking variables from the cancer classification model, shown separately for non-cancer (Y = 0, (**A**)) and cancer (Y = 1, (**B**)) participants. Solid lines denote linear fits, with Pearson correlation coefficients indicated in each panel. Across both outcome groups, correlations were weak and inconsistent in direction, supporting minimal age dependence of individual model features.

**Association of renal function with cancer status**

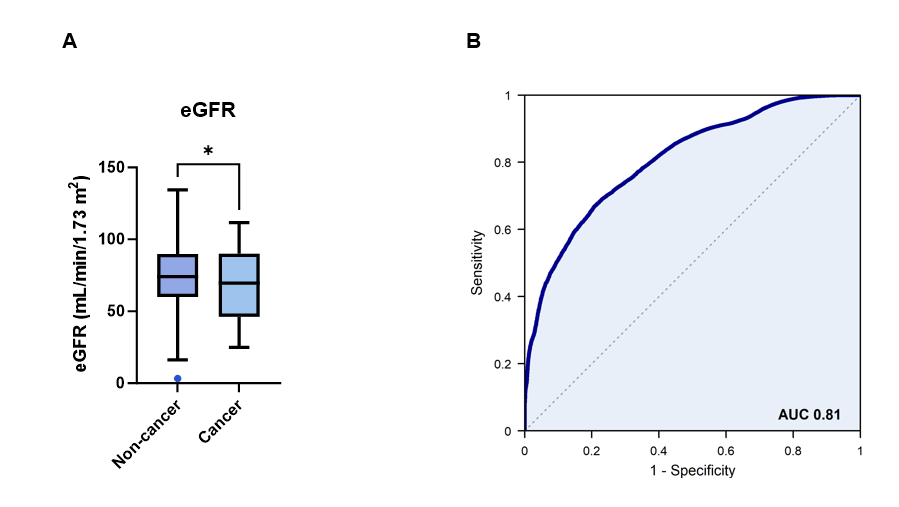

**SI Figure 5** **Association of renal function with cancer status and discriminatory performance.** **(A)** Creatinine-derived estimated glomerular filtration rate (eGFR) in cancer and non-cancer participants (p = 0.013). Boxes indicate median and interquartile range; whiskers denote Tukey limits. **(B)** Receiver operating characteristic (ROC) curves showing model performance with all variables, including eGFR (ROC AUC 0.809 (95% CI 0.803-0.8154)). Despite a modest univariable association, eGFR was not prioritised in multivariable feature selection and did not materially influence model performance.

**Validation in the SCAN1 Cohort**

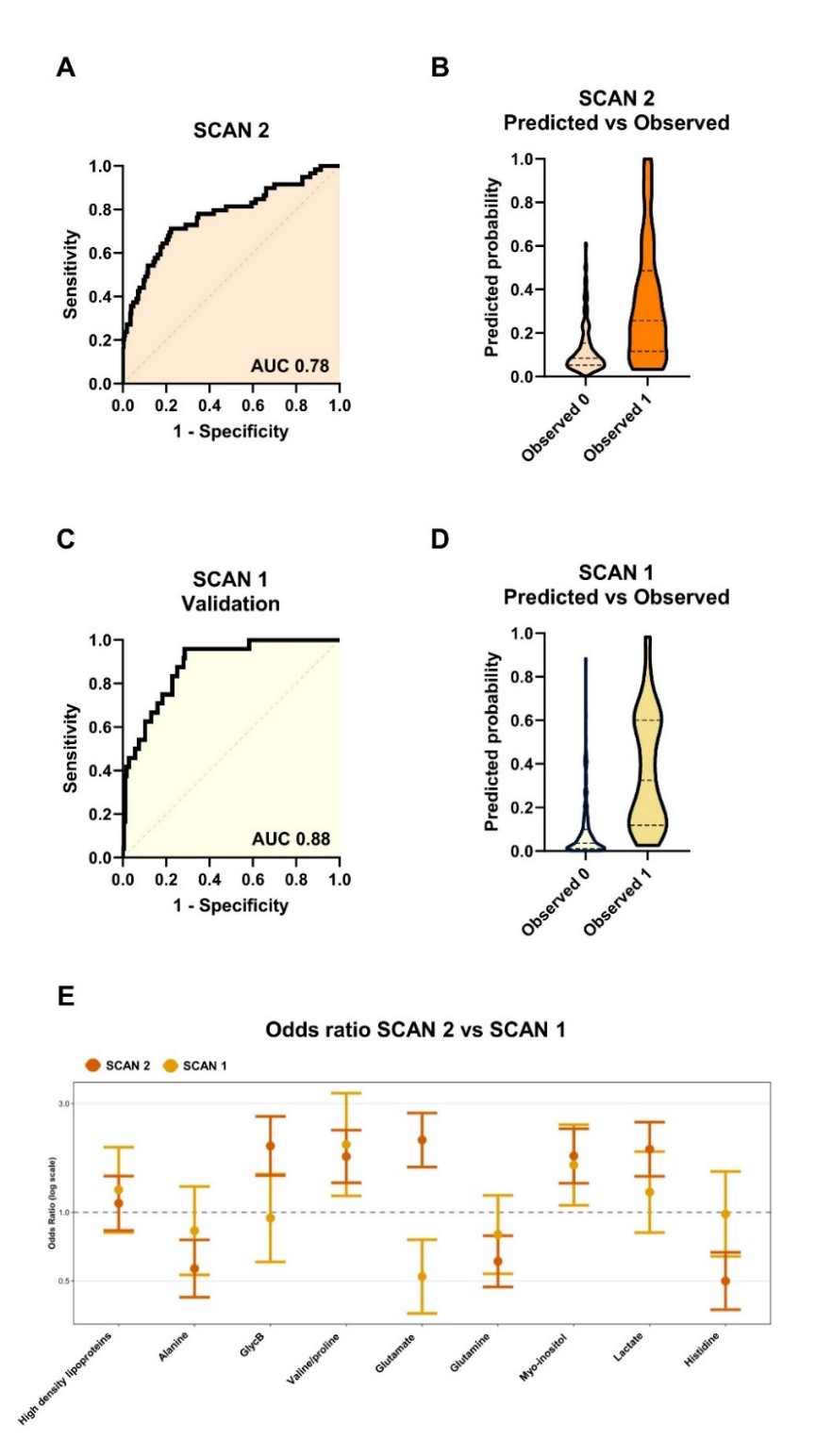

### **SI Figure 6 External validation of the nine most discriminatory NMR-derived metabolites between SCAN2 and SCAN1 samples. (A)** Receiver-operating characteristic (ROC) curve for the SCAN2 model built using the nine top-ranked metabolites (glutamate, histidine, myo-inositol, glycine, valine, leucine, isoleucine, HDL-p, SHDL-p), yielding an AUC of 0.78. **(B)** Predicted versus observed probabilities for cancer and non-cancer cases in SCAN2, showing clear separation between groups. **(C)** ROC curve for validation in SCAN1 using the same nine metabolites, demonstrating higher discrimination (AUC 0.88). **(D)** Predicted versus observed probabilities in SCAN1, confirming reproducible classification performance across cohorts. **(E)** Comparison of odds ratios (log scale, ±95% CI) for the nine metabolites in SCAN2 and SCAN1, showing consistent effect directions: amino-acid metabolites exhibited higher odds of cancer, whereas HDL-related measures were associated with lower odds in both cohorts.

**Discrimination between cancer and non-cancer individuals using NMR metabolomics and LC-MS glycomics**

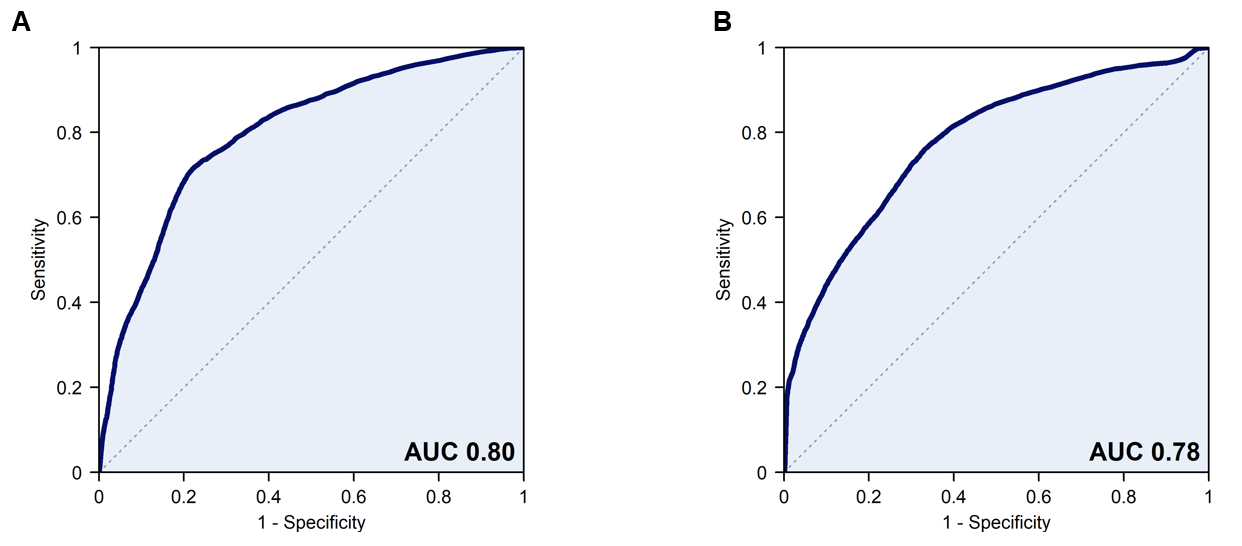

**SI Figure 7 Diagnostic performance of individual analytical platforms for distinguishing cancer from non-cancer in the SCAN2 cohort** (*n* = 369; 59 cancer, 310 non-cancer). **(A)** Receiver-operating characteristic (ROC) curve for the NMR model integrating untargeted metabolomics and targeted AXINON clinical parameters. The model achieved an AUC = 0.799 (95% CI 0.793-0.805), demonstrating strong discriminatory capacity between cancer and non-cancer cases. **(B)** ROC curve for the semi-targeted glycomics model combining fluorescence and MS-based data, which achieved an AUC = 0.778 (95% CI 0.771-0.785), indicating moderate but consistent discrimination across the same cohort. Both models were derived using random forest classifiers with stratified cross-validation and validated in the full SCAN2 dataset.

SI Table 2 Top discriminatory variables distinguishing cancer from non-cancer cases. Variables were selected from the integrated random-forest model including all participants (*n* = 369; 59 cancer, 310 non-cancer) and are ranked by mean decrease Gini index. Odds ratios (OR) with 95% confidence intervals quantify the direction and strength of association with cancer. Statistical significance was assessed using two-sided *t*-tests, with *p*-values corrected for multiple testing using the Benjamini–Hochberg method. Fluorescence-derived glycan clusters correspond to retention-time peaks, with the predominant glycan structure indicated for each cluster.

| **Variable** | **Platform** | **Mean decrease Gini** | **OR** | **Lower**  **95% CI** | **Upper**  **95% CI** | **p-value**  **(raw)** | **p-value**  **(adj.)** |
| --- | --- | --- | --- | --- | --- | --- | --- |
| FA2G2S1 | Glycan (MS) | 2.827 | 0.299 | 0.200 | 0.448 | 4.68E-08 | 7.81E-07 |
| FA2BG1 | Glycan (MS) | 1.876 | 0.514 | 0.386 | 0.685 | 2.35E-04 | 4.15E-04 |
| M5A1G1S1 | Glycan (MS) | 1.570 | 0.406 | 0.298 | 0.553 | 2.28E-06 | 9.77E-06 |
| Glutamate | NMR | 1.424 | 2.075 | 1.579 | 2.726 | 5.40E-07 | 3.63E-06 |
| M4A1G1S1 | Glycan (MS) | 1.400 | 0.433 | 0.320 | 0.586 | 8.01E-06 | 2.40E-05 |
| FA2G2 | Glycan (MS) | 1.170 | 0.379 | 0.278 | 0.516 | 7.26E-07 | 3.63E-06 |
| HDL-p | NMR (AXINON) | 1.115 | 0.435 | 0.328 | 0.576 | 3.35E-07 | 3.35E-06 |
| M5A1G1S1-4 | Glycan (MS) | 1.101 | 0.713 | 0.533 | 0.954 | 8.46E-02 | 9.76E-02 |
| Myo-inositol | NMR | 1.062 | 1.767 | 1.342 | 2.327 | 2.21E-03 | 3.01E-03 |
| SHDL-p | NMR (AXINON) | 1.034 | 0.435 | 0.324 | 0.582 | 5.21E-08 | 7.81E-07 |
| Histidine | NMR | 1.027 | 2.441 | 1.562 | 3.817 | 3.69E-05 | 8.51E-05 |
| M5A1G1 | Glycan (MS) | 1.015 | 0.397 | 0.294 | 0.537 | 6.13E-07 | 3.63E-06 |
| FA2G2S1 | Glycan (MS) | 0.997 | 0.299 | 0.200 | 0.448 | 6.23E-05 | 1.34E-04 |
| FA2G2S2 | Glycan (MS) | 0.977 | 0.564 | 0.439 | 0.724 | 1.55E-04 | 3.10E-04 |
| FA2BG2 | Glycan (MS) | 0.933 | 0.524 | 0.398 | 0.691 | 1.90E-04 | 3.56E-04 |
| Cluster 25.9 min (M5A1G1) | Glycan (fluorescence) | 0.931 | 0.415 | 0.307 | 0.559 | 4.90E-06 | 1.77E-05 |
| Histidine | NMR | 0.930 | 1.905 | 1.377 | 2.635 | 3.77E-04 | 6.28E-04 |
| Cluster 40.2 min (A3G3S2) | Glycan (fluorescence) | 0.904 | 1.653 | 1.235 | 2.213 | 6.16E-03 | 7.70E-03 |
| Mobile -CH_3_ HDL/LDL | NMR | 0.893 | 0.817 | 0.615 | 1.085 | 2.31E-01 | 2.48E-01 |
| glutamine/glutamate | NMR | 0.881 | 2.082 | 1.503 | 2.884 | 8.80E-04 | 1.39E-03 |
| Cluster 40.3 min (A3G3S3) | Glycan (fluorescence) | 0.881 | 2.062 | 1.552 | 2.739 | 1.07E-05 | 2.67E-05 |
| Myo-inositol | NMR | 0.880 | 1.412 | 1.082 | 1.843 | 7.91E-02 | 9.50E-02 |
| FA3G3S3 | Glycan (MS) | 0.872 | 0.432 | 0.313 | 0.595 | 5.30E-06 | 1.77E-05 |
| Cluster 22.4 min (FA1G1) | Glycan (fluorescence) | 0.834 | 0.485 | 0.363 | 0.649 | 1.01E-03 | 1.51E-03 |
| Cluster 23.4 min (FA1G1) | Glycan (fluorescence) | 0.831 | 0.550 | 0.416 | 0.728 | 1.06E-03 | 1.51E-03 |
| Cluster 31.3 min (FA2G2S2) | Glycan (fluorescence) | 0.820 | 0.649 | 0.472 | 0.891 | 1.33E-05 | 1.88E-05 |
| Lymphocyte count | Haematology | 0.818 | 0.739 | 0.552 | 0.990 | 7.14E-01 | 7.14E-01 |
| Glutamine | NMR | 0.817 | 2.014 | 1.534 | 2.644 | 9.52E-06 | 2.60E-05 |
| 3-Hydroxybutyrate | NMR | 0.817 | 1.590 | 1.236 | 2.045 | 2.96E-03 | 3.86E-03 |
| FA1G1S1 | Glycan (MS) | 0.812 | 0.851 | 0.640 | 1.130 | 2.69E-01 | 2.78E-01 |

SI Table 3 Top discriminatory variables distinguishing cancer from non-cancer cases (selected sub-set based on cancer type and comorbidities). Variables were selected from the integrated random-forest model including selected participants (*n* = 309; 32 cancer, 277 non-cancer) and are ranked by mean decrease Gini index. Odds ratios (OR) with 95% confidence intervals quantify the direction and strength of association with cancer. Statistical significance was assessed using two-sided *t*-tests, with *p*-values corrected for multiple testing using the Benjamini–Hochberg method. Fluorescence-derived glycan clusters correspond to retention-time peaks, with the predominant glycan structure indicated for each cluster.

| **Variable** | **Platform** | **Mean Decrease Gini** | **OR** | **Lower 95% ci** | **Upper 95% ci** | **p-value (raw)** | **p-value (adj.)** |
| --- | --- | --- | --- | --- | --- | --- | --- |
| FA2G2S1 | Glycan (MS) | 1.450 | 0.241 | 0.140 | 0.414 | 1.86E-05 | 9.29E-05 |
| M4A1G1S1 | Glycan (MS) | 1.157 | 0.350 | 0.237 | 0.518 | 8.56E-05 | 2.85E-04 |
| M5A1G1S1 | Glycan (MS) | 1.076 | 0.296 | 0.194 | 0.451 | 6.68E-06 | 5.01E-05 |
| Cluster 28.1 min (FA2BG1S1) | Glycan (fluorescence) | 0.909 | 0.390 | 0.263 | 0.580 | 2.87E-04 | 6.15E-04 |
| FA2BG1 | Glycan (MS) | 0.845 | 0.461 | 0.316 | 0.671 | 1.63E-03 | 2.99E-03 |
| Glutamine | NMR | 0.801 | 2.568 | 1.815 | 3.634 | 1.28E-04 | 3.24E-04 |
| Glutamate | NMR | 0.781 | 2.680 | 1.890 | 3.801 | 3.55E-06 | 3.55E-05 |
| FA2G2 | Glycan (MS) | 0.773 | 0.281 | 0.183 | 0.430 | 4.40E-07 | 6.60E-06 |
| Myo-inositol | NMR | 0.727 | 1.323 | 0.992 | 1.765 | 3.06E-01 | 3.40E-01 |
| Glutamine/glutamate | NMR | 0.709 | 2.998 | 1.927 | 4.665 | 2.58E-03 | 4.07E-03 |
| Leucine | AXINON | 0.681 | 0.625 | 0.421 | 0.928 | 3.56E-02 | 4.45E-02 |
| M5A1G1 | Glycan (MS) | 0.668 | 0.332 | 0.227 | 0.488 | 1.82E-05 | 9.29E-05 |
| Cluster 25.9 min (M5A1G1) | Glycan (fluorescence) | 0.664 | 0.290 | 0.192 | 0.440 | 2.51E-07 | 6.60E-06 |
| M5A1G1S1-4 | Glycan (MS) | 0.638 | 0.677 | 0.460 | 0.996 | 2.01E-01 | 2.32E-01 |
| Glutamate | NMR | 0.630 | 2.328 | 1.681 | 3.225 | 1.40E-04 | 3.24E-04 |
| Cluster 40.2 min (A3G3S2) | Glycan (fluorescence) | 0.619 | 1.762 | 1.210 | 2.567 | 3.21E-02 | 4.19E-02 |
| Cluster 35.2 min (complex glycan) | Glycan (fluorescence) | 0.617 | 0.972 | 0.674 | 1.401 | 9.68E-01 | 9.68E-01 |
| Myo-inositol | NMR | 0.538 | 2.509 | 1.612 | 3.905 | 4.46E-03 | 6.37E-03 |
| Formate | NMR | 0.538 | 1.038 | 0.710 | 1.516 | 8.50E-01 | 8.79E-01 |
| Cluster 31.3 min (FA2G2S2) | Glycan (fluorescence) | 0.538 | 0.680 | 0.451 | 1.024 | 4.91E-01 | 5.26E-01 |
| FA1G1S1 | Glycan (MS) | 0.519 | 0.741 | 0.508 | 1.080 | 1.49E-01 | 1.78E-01 |
| Cluster 31.7 min (A2G2S2) | Glycan (fluorescence) | 0.512 | 0.406 | 0.266 | 0.619 | 4.09E-04 | 8.18E-04 |
| Lactate | NMR | 0.491 | 2.176 | 1.553 | 3.048 | 1.69E-03 | 2.99E-03 |
| Cluster 23.4 min (FA1G1) | Glycan (fluorescence) | 0.490 | 0.447 | 0.311 | 0.643 | 1.83E-03 | 3.05E-03 |
| A1G1S1 | Glycan (MS) | 0.489 | 0.552 | 0.376 | 0.811 | 2.10E-02 | 2.86E-02 |
| FA2G2S1 | Glycan (MS) | 0.482 | 0.407 | 0.292 | 0.569 | 1.31E-04 | 3.24E-04 |
| HDL-p | AXINON | 0.467 | 0.421 | 0.300 | 0.589 | 1.16E-04 | 3.24E-04 |
| Valine | AXINON | 0.459 | 0.427 | 0.283 | 0.642 | 6.69E-05 | 2.51E-04 |
| Histidine | NMR | 0.441 | 2.486 | 1.389 | 4.450 | 3.36E-03 | 5.03E-03 |
| Cluster 41.5 min (A4G4S4) | Glycan (fluorescence) | 0.440 | 2.416 | 1.692 | 3.448 | 5.85E-05 | 2.51E-04 |

SI Table 4 Top discriminatory variables distinguishing metastatic from non-metastatic cancer. Variables were selected from the integrated random-forest model including all participants (*n* = 59; 29 metastatic cancer, 30 non-metastatic cancer) and are ranked by mean decrease Gini index. Odds ratios (OR) with 95% confidence intervals quantify the direction and strength of association with cancer. Statistical significance was assessed using two-sided *t*-tests, with *p*-values corrected for multiple testing using the Benjamini–Hochberg method. Fluorescence-derived glycan clusters correspond to retention-time peaks, with the predominant glycan structure indicated for each cluster.

| **Variable** | **Platform** | **Mean decrease Gini** | **OR** | **Lower**  **95% CI** | **Upper**  **95% CI** | **p-value**  **(raw)** | **p-value**  **(adj.)** |
| --- | --- | --- | --- | --- | --- | --- | --- |
| Histidine | NMR | 0.629 | 0.413 | 0.211 | 0.809 | 0.004 | 0.018 |
| FA4G4S4 | Glycan (MS) | 0.521 | 2.325 | 1.250 | 4.327 | 0.004 | 0.018 |
| Histidine | NMR | 0.481 | 0.300 | 0.127 | 0.713 | 0.002 | 0.015 |
| Alanine | NMR | 0.480 | 0.387 | 0.195 | 0.767 | 0.002 | 0.015 |
| Alanine | NMR | 0.474 | 0.384 | 0.193 | 0.764 | 0.002 | 0.015 |
| Cluster 31.7 min | Glycan (fluorescence) | 0.429 | 0.287 | 0.119 | 0.695 | 0.002 | 0.015 |
| Alanine | NMR (AXINON) | 0.403 | 0.362 | 0.180 | 0.729 | 0.002 | 0.015 |
| Myo-inositol | NMR | 0.394 | 1.399 | 0.774 | 2.531 | 0.270 | 0.338 |
| Cluster 40.8 min | Glycan (fluorescence) | 0.372 | 1.940 | 1.064 | 3.539 | 0.022 | 0.050 |
| Cluster 33.9 min | Glycan (fluorescence) | 0.333 | 0.482 | 0.260 | 0.894 | 0.015 | 0.042 |
| Myo-inositol | NMR | 0.322 | 1.241 | 0.710 | 2.168 | 0.472 | 0.488 |
| Free choline/-N(CH3)3 | NMR | 0.291 | 0.668 | 0.388 | 1.150 | 0.153 | 0.219 |
| Glucose region | NMR | 0.281 | 1.430 | 0.805 | 2.539 | 0.222 | 0.290 |
| Myo-inositol | NMR | 0.276 | 1.529 | 0.811 | 2.882 | 0.176 | 0.240 |
| Cluster 23.8 min | Glycan (fluorescence) | 0.274 | 0.584 | 0.328 | 1.038 | 0.068 | 0.128 |
| Cluster 24.4 min | Glycan (fluorescence) | 0.265 | 0.472 | 0.256 | 0.869 | 0.010 | 0.030 |
| FA4G4S4 | Glycan (MS) | 0.260 | 4.200 | 1.383 | 12.758 | 0.007 | 0.024 |
| Isoleucine | NMR | 0.254 | 0.653 | 0.370 | 1.150 | 0.137 | 0.216 |
| Mobile -CH3 HDLs | NMR | 0.252 | 1.498 | 0.855 | 2.623 | 0.154 | 0.219 |
| Myo-inositol | NMR | 0.251 | 1.365 | 0.754 | 2.470 | 0.307 | 0.368 |
| Glutamine | NMR | 0.248 | 0.595 | 0.340 | 1.040 | 0.060 | 0.121 |
| FA3G3S1 | Glycan (MS) | 0.247 | 1.632 | 0.915 | 2.909 | 0.087 | 0.154 |
| A4G4S2 | Glycan (MS) | 0.242 | 1.203 | 0.713 | 2.030 | 0.574 | 0.574 |
| Cluster 25.9 min | Glycan (fluorescence) | 0.235 | 0.436 | 0.227 | 0.838 | 0.008 | 0.027 |
| Myo-inositol | NMR | 0.234 | 1.275 | 0.723 | 2.247 | 0.408 | 0.447 |
| M4A1G1S1 | Glycan (MS) | 0.230 | 1.226 | 0.727 | 2.067 | 0.417 | 0.447 |
| Cluster 39.0 min | Glycan (fluorescence) | 0.228 | 1.741 | 0.902 | 3.361 | 0.095 | 0.159 |
| Citrate | NMR | 0.225 | 1.256 | 0.744 | 2.120 | 0.382 | 0.441 |
| FA4FG4S4 | Glycan (MS) | 0.224 | 2.058 | 1.129 | 3.753 | 0.018 | 0.045 |
| Glucose region | NMR | 0.221 | 0.571 | 0.314 | 1.040 | 0.057 | 0.121 |
